# The assembly of neutrophil inflammasomes during COVID-19 is mediated by type I interferons

**DOI:** 10.1101/2023.09.07.23295190

**Authors:** Luz E. Cabrera, Suvi T. Jokiranta, Sanna Mäki, Simo Miettinen, Ravi Kant, Lauri Kareinen, Tarja Sironen, Jukka-Pekka Pietilä, Anu Kantele, Eliisa Kekäläinen, Hanna Lindgren, Pirkko Mattila, Anja Kipar, Olli Vapalahti, Tomas Strandin

## Abstract

The severity of COVID-19 is linked to excessive inflammation. Neutrophils represent a critical arm of the innate immune response and are major mediators of inflammation, but their role in COVID-19 pathophysiology remains poorly understood. We conducted transcriptomic profiling of neutrophils obtained from patients with mild and severe COVID-19, as well as from SARS-CoV-2 infected mice, in comparison to non-infected healthy controls. In addition, we investigated the inflammasome formation potential in neutrophils from patients and mice upon SARS-CoV-2 infection. Transcriptomic analysis of polymorphonuclear cells (PMNs), consisting mainly of mature neutrophils, revealed a striking type I interferon (IFN-I) gene signature in severe COVID-19 patients, contrasting with mild COVID-19 and healthy controls. Notably, low-density granulocytes (LDGs) from severe COVID-19 patients exhibited an immature neutrophil phenotype and lacked this IFN-I signature. Moreover, PMNs from severe COVID-19 patients showed heightened nigericin-induced caspase1 activation, but reduced responsiveness to exogenous inflammasome priming. Furthermore, IFN-I emerged as a priming stimulus for neutrophil inflammasomes, which was confirmed in a COVID-19 mouse model. These findings underscore the crucial role of neutrophil inflammasomes in driving inflammation during severe COVID-19. Altogether, these findings open promising avenues for targeted therapeutic interventions to mitigate the pathological processes associated with the disease.

## Introduction

Severe COVID-19 is characterized by a dysregulated immune response with an excessive production of pro-inflammatory cytokines and chemokines. Type I interferons (IFN-I) are critical antiviral cytokines in the innate immune responses against viral infections, drawing particular attention amidst the COVID-19 pandemic (1–3). While the IFN-I response helps to limit virus replication (3), its prolonged and uncontrolled activation is detrimental to the overall health of the patient (4). As part of the pro-inflammatory response, neutrophils are rapidly recruited to the site of infection in response to SARS-CoV-2 infection (5, 6). Prominent neutrophil recruitment in severe COVID-19 is associated with an increased number of immature low-density granulocytes (LDGs) in the circulation (7–9). The increased production and subsequent early release of immature cells from the bone marrow occurs in response to emergency myelopoiesis (9). This process is initiated by the body to enable the recruitment of innate immune cells into the tissues and to replenish the depleted leukocyte pool, in an effort to combat viral infections including SARS-CoV-2 (10). However, the premature release of these cells could be associated with the increased degranulation and formation of neutrophil extracellular traps (NETs) reported during SARS-CoV-2 infection, to which LDGs have a higher propensity than polymorphonuclear cells (PMN) (5, 6, 11).

Neutrophils are involved in several aspects of inflammatory processes, including the release of reactive oxygen species (ROS) and other pro-inflammatory mediators such as Interleukin-6 (IL-6) and IL-8. In addition, recent reports on COVID-19 highlight that neutrophils could be a major source of inflammasome derived IL-1β, which has been implicated as a substantial contributor to COVID-19 pneumonia (12). Inflammasomes are intracellular multiprotein complexes involved in the inflammatory response. In the presence of a pathogen, antigen recognition by the immune system triggers the assembly of the inflammasome, a step known as the first signal. This is followed by the recruitment of adaptor molecules that activate NOD-like receptor (NLR) family members and the binding of the apoptosis-associated speck-like protein (ASC), finally activating the inflammasome complex (13). The triggered assembly of this complex is known as the second signal. Studies have shown that SARS-CoV-2 infection induces significant inflammasome activation in circulating and lung-infiltrating myeloid cells, such as monocytes and neutrophils (14–17). However, while the precise mechanism by which inflammasomes are activated in monocytes/macrophages is well established, less is known about molecular mechanisms of inflammasome formation in neutrophils. Thus, this study investigates the inflammasome formation in neutrophils during COVID-19 in more detail, also focusing on the different developmental stages of these cells. In addition, a recently established COVID-19 mouse model served to further explore the role of IFN-I in neutrophil inflammasome assembly.

## Materials and methods

### Patient population

Adult clinical patients with confirmed COVID-19 (RT-PCR positive for SARS-CoV-2) at Helsinki University Hospital (HUH) (hospitalized: n = 34; outpatients: n = 8) were enrolled in the present study. Blood samples were collected during hospitalization for the severe COVID-19 group, and after confirmation of diagnosis for the mild COVID-19 outpatient group. Samples for RNA sequencing were collected in 2020 and representing infections by the original and early SARS-CoV-2 variants, whereas samples for *ex vivo* culture experiments were collected in 2021-2022 likely representing infections by omicron subvariants of SARS-CoV-2. As controls, healthy blood donors were included for RNA sequencing (n = 7, age 57 ± 7, male/female 3/4) and *ex vivo* culturing experiments (n = 9, age 38 ± 14, male/female 4/5). The study was approved by the Ethics Committee of the Hospital District of Helsinki and Uusimaa (HUS/853/2020, HUS/1238/2020). All volunteers gave a written informed consent, in accordance with the Declaration of Helsinki. For clinical correlation analysis, severe COVID-19 patients were further categorized by severity based on their need for hospitalization and oxygen supplementation, as described previously (7). For each patient, medical history and clinical data were collected through retrospective patient record review and are presented for the severe COVID-19, hospitalized patients in Table 1 and as previously described (7). Calprotectin was measured from serum (diluted 1:1000) by ELISA, according to the manufacturer’s protocol (calprotectin/S100A8 DuoSet kit, R&D systems).

**Table 1.** Clinical parameters of hospitalized patients (n=34).

| Parameter | Range (Mean ± SD) or ratio |
| --- | --- |
| Age (years) | 22 - 80 (58.41 ± 13.63) |
| Gender (male:female) | 23:11 |
| Hospitalization (days) | 3 - 38 (12 ± 8.63) |
| WHO Ordinal scale for Clinical improvement: Baseline | 1 - 6 (4 ± 1.07) |
| WHO Ordinal scale for Clinical improvement: Worst | 2 - 8 (4 ± 1.42) |
| Deaths (count/total) | 1/34 |
| ICU (count/total) | 9/34 |
| Corticosteroid use (count/total) | 13/34 |
| Length of stay in ICU (days) | 1 - 27 (7.38 ± 9.49) |
| Max CRP (mg/L) | 6 - 466 (159.56 ± 104.06) |
| Min lymphocyte count (E<sup>9</sup>/L) | 0.08 - 2.02 (0.79 ± 0.42) |
| Max neutrophil count (E<sup>9</sup>/L) | 1.08 - 29.86 (7.85 ± 5.73) |
| Sampling day neutrophil count (E<sup>9</sup>/L) | 0.64 - 20.52 (5.65 ± 3.95) |

The World Health Organization (WHO) Ordinal Scale for clinical improvement is a tool designed specifically to assess and measure the progression and clinical improvement of patients (65). COVID-19 scoring: 1= no limitations of activity, 2= limitations of activity, 3= no oxygen therapy, 4= oxygen by mask or nasal cannulae, 5= non-invasive ventilation or high-flow oxygen, 6= invasive mechanical ventilation without other organ support, 7= invasive mechanical ventilation with other organ support, 8= dead. The baseline score represents the timepoint of the first laboratory sample taken, serving as a reference point for measuring improvement and establishing a starting point for comparison. In contrast, the worst score represents the most severe or critical state of the disease. None of the patients whose samples were used for RNA-seq underwent corticosteroid treatment. *Abbreviations: WHO= World Health Association. ICU = Intensive Care Unit. CRP = C reactive protein. Min = minimum. Max = maximum.*

### Isolation of granulocytes from human blood

Blood samples from COVID-19 patients and healthy controls (HC) were collected in EDTA vacutainer tubes and transported to the laboratory. Peripheral blood mononuclear cells (PBMCs) or polymorphonuclear cells (PMNs) were isolated from whole blood by density gradient centrifugation using either Ficoll-Paque Plus (GE Healthcare) or Polymorphprep (Axis-Shield) respectively, following standard procedures including the use of 2 mM EDTA in PBS and red blood cell lysis with ACK lysis buffer (Lonza by Thermo Fisher). Subsequently, isolation of CD66^+^ granulocytes (low-density granulocytes, LDGs) from the PBMC fraction was performed using the CD66abce MicroBead Kit (Miltenyi Biotec, Germany) with an MS column, according to the manufacturer’s instructions. Both the positively selected CD66^+^ LDGs and the isolated PMNs were then washed and counted, using a TC20^TM^ Automated Cell Counter (Bio-Rad Laboratories, Inc.) with trypan blue staining for dead cell exclusion. All described procedures in this section were done at room temperature. An aliquot of cells was lysed in Trizol reagent (Thermo Fisher Scientific, USA) and stored at –80°C for later extraction of total RNA and subsequent RNA sequencing (RNA-seq) analysis.

### Caspase1 activity

Caspase1 activity was assessed in isolated cells after 2 h of culture (1 million cells/ml) using the caspase-Glo^®^1 inflammasome assay (Promega) according to the manufacturer’s protocol, with 2.5 µM nigericin (Invivogen) treatment as the activator. The resulting luminescence was measured by a Hidex Sense microplate reader (Hidex).

### Soluble factor stimulation assays

Isolated granulocytes from HC and COVID-19 patients were cultured at 2 million cells/ml in RPMI 1640 supplemented with 10% fetal bovine serum (R10) at 37°C. Cells were primed (1^st^ signal) with either LPS (20 ng/ml, Sigma Aldrich) or IFN-I (combination of 2.7*10^4^ IU/ml IFN-α and IFN-β, Immunotools) for 4 h, followed by activation (2^nd^ signal) by 2.5 µM nigericin or monosodium urate crystals (MSU, 100 µg/ml, Invivogen) for an additional 4 h. For the 24 h stimulation experiments, nigericin was added to the cultured cells, in the presence or absence of inflammasome inhibitors MCC995 (2 µg/ml) and Ac-YVAD-FMK (20 µg/ml, both from Invivogen). Cells were pelleted by centrifugation at 400 G for 5 min and stored in Trizol at – 80°C for later RNA extraction whereas supernatants were used to measure IL-1β, IL-18, myeloperoxidase (MPO) and IL-8 by ELISAs according to the manufacturer’s protocols (DuoSet kits from R&D systems). LDH was measured in supernatants using Cyquant LDH cytotoxicity assay (ThermoFisher). HL-60 cells (ATCC #CCL-240) were activated similarly to neutrophils after a 5-day differentiation period induced by 1 % DMSO.

### Virus propagation

The SARS-CoV-2 hCoV-19/Finland/THL-202117309/2021 (delta strain B.1.617.2) and the mouse-adapted strain MaVie (18) were propagated in VeroE6-TMPRSS2 cells (kidney epithelial cells expressing the transmembrane protease serine 2) (19) grown in DMEM supplemented with 10% inactivated FCS, 100 IU/mL Penicillin, 100 μg/mL Streptomycin and 2 mM L-glutamine at 37°C. The virus was purified from supernatants by ultracentrifugation (SW28 rotor, 27,000 rpm, 90 min, +4 °C) through a 0.22 μm-filtered 30% ultra-pure sucrose cushion (in PBS), to obtain virus preparations free of cell culture contaminants. Virus titers were calculated by the median tissue culture infectious dose (TCID50) after assessing cytopathic effects by crystal violet staining of cell cultures infected for 5 days with serially diluted virus.

### RNA sequencing

Neutrophils isolated from different cohorts comprised three PMN groups (severe COVID-19, mild COVID-19, and healthy controls), and one LDG group (given that these cells were rare in mild COVID-19 patients and HC, only LDGs from patients with severe COVID-19 were included).

cDNA synthesis from total RNA was performed according to Takara SMARTseq v4 Ultra-low input RNA kit for Sequencing user manual (Takara Bio, Mountain View, CA, USA) followed by Illumina Nextera XT Library preparation according to Illumina Nextera XT Reference Guide (Illumina, San Diego, CA, USA). UDI index setup was used for the Nextera XT libraries. Library quality check was performed using LabChip GX Touch HT High Sensitivity assay (PerkinElmer, USA) and libraries were pooled based on the concentrations acquired from the assay. The pooled libraries were quantified for sequencing using KAPA Library Quantification Kit (KAPA Biosystems, Wilmington, MA, USA) and sequenced on the Illumina NovaSeq6000 system for 200 cycles using S1 flow cell (Illumina, San Diego, CA, USA). Read length for the paired-end run was 2×101 bp.

### RNA data analysis

Principal Component Analysis (PCA) and enrichment analyses were obtained using ExpressAnalyst (20). Briefly, PCA was performed to identify patterns in the data and reduce the dimensionality of the dataset, where the top principal components were selected based on the percentage of variance explained. For enrichment analyses, Gene Set Enrichment Analysis (GSEA) and Over-Representation Analysis (ORA) were performed on the top 5000 DE genes identified by DESeq2 (adjusted P value < 0.05, log2FC >1) (20). GSEA was used to identify enriched signaling pathways using the Reactome database, while ORA was used to identify enriched pathways using the KEGG database. The resulting p-values were corrected for multiple testing using the Benjamini-Hochberg method, and pathways with a corrected p-value <0.05 were considered significant.

To visualize the expression patterns of the differentially expressed (DE) genes, the data was analyzed using the AltAnalyze software (21), which selected the top 118 genes based on correlation and determined the heatmap clustering, using the Euclidean distance metric and the complete linkage method. Then, the obtained heatmap was re-generated using heatmapper.ca (22) for better visualization.

CIBERSORTx, a machine learning algorithm that infers cell type proportions using a reference gene expression matrix of known cell types (25) was used to perform RNA-seq deconvolution on the gene expression data to estimate the abundance of immune cell types in the samples (23). The signature matrix used was taken from Lasalle *et al.* (8). This reference matrix made use of a published whole-blood single-cell dataset (9), and included the main immune cell types: monocytes, NK cells, T lymphocytes, B lymphocytes, plasmablasts and neutrophils, the latter subclassified into mature and immature. The smaller subsets of granulocytes (eosinophils and basophils) are not considered separately and are most likely categorized as neutrophils in the bulk data deconvolution. Nonetheless, the resulting cell type proportions were used to compare the immune cell composition between groups.

Additionally, the determination of sample purity (>65% identified as neutrophils) served as a limiting parameter for the visualization of differentially expressed inflammasome related genes from the RNA sequencing results, which were selected and graphed in a heatmap using heatmapper.ca (22), clustered by complete linkage and ordered by Spearman’s rank.

### Volcano Plots

To visualize differentially expressed (DE) genes between groups from human and mice RNA-seq results previously identified by DESeq2, a volcano plot was generated using GraphPad Prism. Genes with a P-adjusted value (padj or FDR) <0.05 were considered significant. Similarly, RNA sequencing data from GSE93996 (24) was reanalyzed, and all DE genes in ATRA-differentiated HL-60 cells were visualized in a volcano plot.

### Single cell transcriptomics data analysis

This study made use of the “COVID-19 Immune Atlas: integration of 5 public COVID-19 PBMC single-cell datasets” available online (25). This standardized data collection contains cells from different assays (10x 3’ v2, 10x 3’ v3, 10x technology and Seq-Well) and consists of a total of 239,696 cells from the peripheral blood, 3,693 of which are neutrophils. These neutrophils were further subclassified as mature (59%) and immature (41%), based on the immune atlas predetermined cell classes. This was confirmed by a CD16b expression in mature neutrophils, and a higher CD66b expression in the immature population. This data was obtained from and analyzed in the Chan Zuckerberg CELLxGENE platform (25).

### Reverse transcription and quantitative PCR (RT-qPCR) for human selected human genes

Total RNA was extracted from unstimulated or *ex vivo* stimulated PMNs using the Trizol reagent (Invitrogen, USA) according to the manufacturer’s protocol. Subsequently, cDNA synthesis was performed using the RevertAid RT Reverse Transcription Kit (Thermo Scientific, USA) as per the manufacturer’s instructions. Quantitative PCR (qPCR) was performed using the Stratagene model (Agilent Technologies) and SYBR Green/ROX master mix (Thermo Scientific, USA). The primer sequences for qPCR are presented in Supplementary Table S1.

Primer specificity was confirmed using melting curve analysis and dissociation curves. The relative expression levels of the genes of interest were calculated using the 2-ΔΔCT method and normalized to the expression of the housekeeping gene GAPDH. Baseline gene expressions of unstimulated samples were statistically assessed using the Mann-Whitney test, while the two-way ANOVA Tukey’s multiple comparisons test was performed for the *ex vivo* stimulated samples.

### Mouse infections

Experimental procedures were approved by the Animal Experimental Board of Finland (license number ESAVI/28687/2020). Female BALB/c mice (Envigo, Indianapolis, IN, USA; 7 to 8 weeks, n = 36 in total) were transferred to the University of Helsinki biosafety level-3 (BSL-3) facility and acclimatized to individually ventilated biocontainment cages (ISOcage; Scanbur, Karl Sloanestran, Denmark) for 7 days with *ad libitum* water and food (rodent pellets). For subsequent experimental infection, the mice were placed under isoflurane anesthesia and inoculated intranasally with 50 µL of SARS-CoV-2 MaVie strain (5*10^5^ TCID50/animal) or PBS (mock-infected control). Daily weighting of all mice was performed, and their well-being was carefully monitored for signs of illness (e.g., changes in posture or behavior, rough coat, apathy, ataxia). Euthanasia was performed by cervical dislocation under terminal isoflurane anesthesia. All animals were dissected immediately after euthanasia, and the lungs were sampled for multiple downstream analyses. The infections were performed as 3 separate experiments (exp) with 12 mice each: 1) Exp 1 included 8 mice infected with MaVie and 4 mock infected mice. At 2 days post infection (dpi), 4 infected and the mock infected mice were euthanized; the remaining infected mice were euthanized at 4 dpi. The right lung was sampled for virus-specific RT-qPCR (1/5) and neutrophil isolation (4/5), the left lung was fixed for histological and immunohistochemical examination. 2) Exp 2 included 8 infected and 4 mock infected mice of which half were euthanized at 2 dpi and 4 dpi, respectively. From these mice, both lung lobes were subjected to neutrophil isolation. 3) Exp 3 included 8 mice that were infected and immediately inoculated intraperitoneally with 250 µg of anti-mouse IFNAR-1 (n = 4) or IgG1 isotype control (n = 4) (Bio-X-Cell, New Hampshire, USA), and 4 mock-infected animals. All mice were euthanized at 2 dpi. Each 1 1/5 of the left lobe was processed for virus-specific RT-qPCR and histology and immunohistochemistry respectively. The remaining approx. 80% of the lungs served for neutrophil isolation.

### Neutrophil isolation from mouse lungs

Neutrophil isolation was performed from the lungs of all mice. The dissected lung tissue was chopped into small pieces using scissors and enzymatically digested with a cocktail of Liberase (50 ug/ml; Roche #05401020001 from Merck) and DnaseI (100 ug/ml; Roche #11284932001 from Merck) in RPMI-1640 for 30 min at 37 °C. The resulting homogenate was diluted 10-fold in R10 and passed through a 70 µm Cell strainer (Pluriselect) to obtain a single-cell suspension. Neutrophils were isolated by positive selection using Ly6G-binding magnetic beads and MS columns according to the manufacturer’s recommendations (Miltenyi Biotec). Neutrophils were isolated with a purity exceeding 95% based on flow cytometry analysis of Ly6G expression.

### RNA sequencing of mouse neutrophils

Mouse neutrophils were isolated, lysed in Trizol (Thermo Scientific) and the RNA extracted in the liquid phase using chloroform. RNA isolation was carried out using the Rneasy micro kit (Qiagen). Isolated RNA (1 ng) underwent whole transcriptome sequencing with ribodepletion. Briefly, RNA sequencing was performed using the Illumina Stranded with RiboZero library preparation method. Sample quality and integrity were assessed using TapeStation RNA analysis. Sequencing was conducted on the Illumina NextSeq platform, followed by standard bioinformatics analysis for gene expression quantification.

The service was provided by the Biomedicum Functional Genomics Unit at the Helsinki Institute of Life Science and Biocenter Finland at the University of Helsinki.

### RT-qPCR of mouse samples

RNA was extracted from dissected lung samples (1/10 of the whole lung) of mice in Exp 1 and Exp 3 as well as isolated neutrophils using Trizol (Thermo Scientific) following the manufacturers’ instructions. The isolated RNA was directly subjected to one-step RT-qPCR analysis based on a previously described protocol using primer-probe sets detecting the viral genome encoding for the RNA-dependent RNA polymerase (RdRp) (26), subgenomic E (27) as well as mouse caspase1, IL1b and GAPDH (Applied biosystems #Mm00438023_m1, #Mm00434228_m1 and #Mm99999915_g1 respectively, Thermo scientific). The PCRs were performed with TaqPath 1-step master mix (ThermoFisher Scientific) using AriaMx instrumentation (Agilent, Santa Clara, CA, USA).

### Histology and immunohistochemistry

From animals in Exp 1 and Exp 3 the whole left lung (Exp 1) or 1/5 of the left lung (Exp 3) were trimmed for histological examination and routinely paraffin wax embedded. Consecutive sections (3 µm) were prepared and routinely stained with hematoxylin-eosin (HE) or subjected to immunohistochemistry (IHC) for the detection of SARS-CoV-2 nucleoprotein (NP) (28) and Ly6G (neutrophil marker); for Exp 3, a further section of the infected lungs was stained for histone H3 (NET marker) (29). All stains followed previously published protocols (30).

### Morphometric analyses

For quantification of SARS-CoV-2 antigen expression and the extent of neutrophil influx into the lungs, a morphometric analysis was undertaken on the slides stained for SARS-CoV-2 NP and Ly6G, respectively. The stained slides were scanned using NanoZoomer 2.0-HT (Hamamatsu, Hamamatsu City, Japan), and several sections of the lung of each animal were quantitatively analysed using the Visiopharm 2022.01.3.12053 software (Visiopharm, Hoersholm, Denmark). The average total tissue area used for quantification was 19.5 ± 6 mm^2^. The morphometric analysis served to quantify the area, in all lung sections of an animal, that showed immunostaining for viral NP and Ly6G, respectively. In Visiopharm, for each section, the lung was manually outlined and annotated as a Region Of Interest (ROI), manually excluding artifactually altered areas. The manual tissue selection was further refined with an Analysis Protocol Package (APP) based on a Decision Forest classifier, with the pixels from the ROI being ultimately classified as either “Tissue” or “Background”. This new “Tissue” ROI, regrouping the different lung samples analysed for each animal, was further quantified by executing two APPs successively. The first APP was based on a Threshold classifier and served to detect and outline areas with immunostaining. The second APP then measured both the surface of the immunostained area (µm^2^) and the surface of the “Tissue” ROI (µm^2^). The percentage of immunostained area (%), expressed as the ratio between the immunostained area and the total area, was obtained for each animal in Excel (Microsoft Office 2019; Microsoft, Redmond, Washington, United States), according to the following formula: ([positive area (µm2)]/ [total area (µm2)]) x 100.

### Statistical analyses

Statistical analysis was performed using GraphPad Prism 8.3 software (GraphPad Software, San Diego, CA, USA) and R software v3.6.3 (R core team). Statistically significant correlations between parameters were assessed by calculating Spearman’s correlation coefficients, and differences between groups were assessed with Mann-Whitney, Kruskall-Wallis or ordinary one-way or 2-way ANOVA tests, depending on sample distribution and the number of groups analyzed. To elaborate, nonparametric tests like Mann-Whitney and Kruskall-Wallis were employed when the data violated assumptions of normality, while ANOVA tests were applied when the data met parametric assumptions.

## Results

### Unsupervised RNA-seq analysis reveals an antiviral gene expression signature of circulating neutrophils in COVID-19 that is strongly influenced by maturity

With our recent findings on increased frequencies of low-density granulocytes (LDGs, isolated from the PBMC fraction) during COVID-19 and their likely relevant role in disease progression (7), we sought to understand in more detail how the transcriptomic profile of LDGs differs from their higher “normal” density counterpart, the circulating polymorphonuclear cells (PMNs) (31), typically consisting mainly of mature neutrophils. Neutrophils isolated from different cohorts comprised three PMN groups (severe COVID-19, mild COVID-19, and healthy controls), and one LDG group. Initial deconvolution of the RNA sequencing (RNA-seq) data allowed us to gain a comprehensive understanding of the cellular composition within PMN and LDG fractions and verified that most cells present in the samples were neutrophils (Supplementary Figure 1A). This analysis also demonstrated that cells in the LDG fraction were predominantly immature neutrophils, meanwhile PMNs were composed of mainly mature neutrophils.

The samples with predominant neutrophil cell populations were selected for subsequent gene expression analysis (neutrophils ≥ 65 %). The high variance in gene expression between PMNs and LDGs was confirmed by principal component analysis (PCA) (Figure 1A), which revealed that the gene expression patterns of COVID-19 LDGs differed from those of all PMNs regardless of the patients’ disease state. Functional enrichment analyses through gene overrepresentation (ORA) and gene-set enrichment analyses (GSEA) (Figure 1B) compared PMNs with LDGs from severe COVID-19 patients. The most statistically significant result was an overrepresentation of the NOD-like receptor signaling pathway in PMNs in contrast with LDGs, highlighting that the different neutrophil fractions have a distinct inflammatory profile.

**Figure 1.**
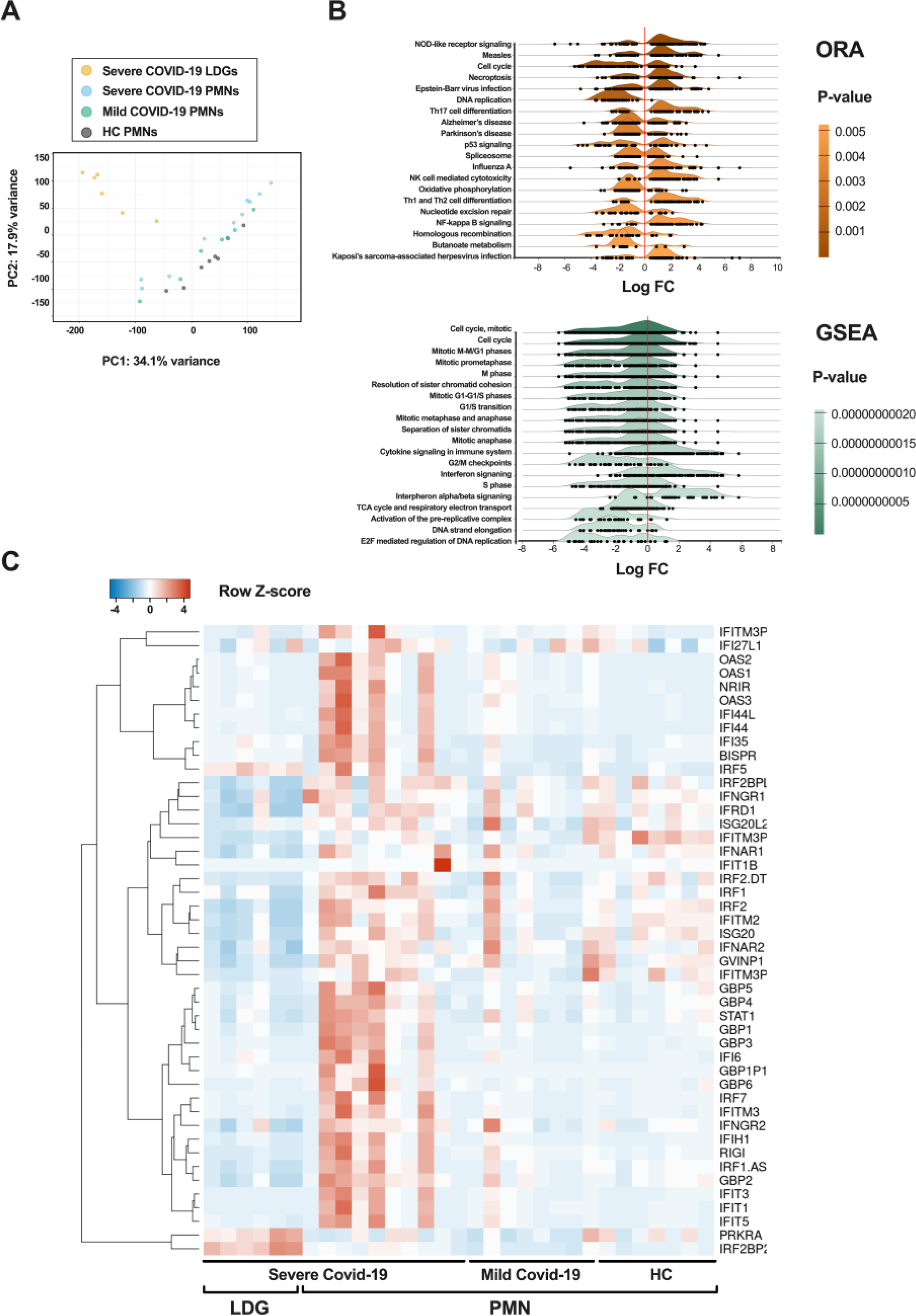
Increased IFN-I related gene expression in mature COVID-19 neutrophils. The analysis was reduced to include only the samples with the highest purity (cell fraction over 0.65 of neutrophils), as identified by CIBERSORTx. (**A**) Principal component analysis (PCA) of the RNA-seq samples (n = 7 PMNs from HC, n = 10 PMNs from severe COVID-19, n=8 PMNs from mild COVID-19, and n = 6 LDGs from severe COVID-19). (**B**) Ridgeline diagrams depicting the top 20 enriched signal pathways from the genes differentially expressed by PMNs versus LDGs during severe COVID-19: overrepresentation analysis (ORA) using KEGG database and gene-set enrichment analysis (GSEA) according to Reactome database. Both enrichment analyses were made using ExpressAnalyst and are sorted by P-value, obtained from Welch’s t-test. (**C**) Heatmap of differentially expressed IFN-related genes in COVID-19 PMNs and LDGs as compared to HC PMNs. RNA sequencing was performed on purified PMNs from healthy controls, mild COVID-19 and severe COVID-19, as well as LDGs from severe COVID-19. The heatmap was clustered by complete linkage and ordered by Spearman’s rank. *FC = fold change*.

This was supported by GSEA, where the most obvious increases in fold changes were the enrichment of the interferon signaling pathways. Another relevant difference was the cell cycle and DNA replication pathways, identified by both ORA and GSEA, which supported our previous findings suggesting LDGs to be predominantly immature cells (7). Furthermore, a heatmap of selected type I IFN (IFN-I) related genes confirmed a robust IFN-I gene signature in severe COVID-19 PMNs, while LDGs from severe COVID-19 distinctively lacked this signature (Figure 1C). Unsupervised clustering analysis, namely Iterative Clustering and Guide Gene Selection (ICGS) using the AltAnalyze software, supported these findings by identifying the top 118 differentially expressed (DE) genes, including several IFN-related genes (Supplementary Fig. 1B). Similarly to the selected samples included in Figure 1, this analysis classified the samples into two major clusters: a first one containing all isolated LDG samples, and a second one comprising all isolated PMN samples. The former cluster consisted of neutrophil antimicrobial and granule marker genes (e.g. *DEFA3, DEFA4, SERPINB10, CTSG*), while in the latter cluster the most significantly upregulated genes in the PMNs from severe COVID-19 subgroup were mainly interferon inducible (e.g. *IFI44L, IFI6, GBP3, IRF7*). These differences were supported by a detailed gene analysis (Supplementary Fig. 2A).

### Inflammasomes are activated in severe COVID-19 PMNs, but not directly by SARS-CoV-2

Looking more closely into PMN fractions, pathway analyses identified the inflammasome related NOD-like and RIG-like receptor signaling pathways among the most significantly overrepresented pathways, differentially expressed in severe COVID-19 PMNs versus HC PMNs (Figure 2A and Supplementary Fig. 2B-C) or mild COVID-19 PMNs (Figure 2B and Supplementary Fig. 2D, E). However, mild COVID-19 PMNs did not significantly differ from HC PMNs in their inflammatory profile (Supplementary Fig. 2F). The increased expression of selected IFN-I (*OAS1, OAS2,* and *IFIT1*) and inflammasome related genes (*CASP1, CASP5, NLRC5* and *NAIP*) was confirmed by RT-qPCR. However, some inflammasome related genes (*IL-1β, NLRP3* and *NLRC4*) were seemingly downregulated, although not statistically significant (Supplementary Fig. 3).

**Figure 2.**
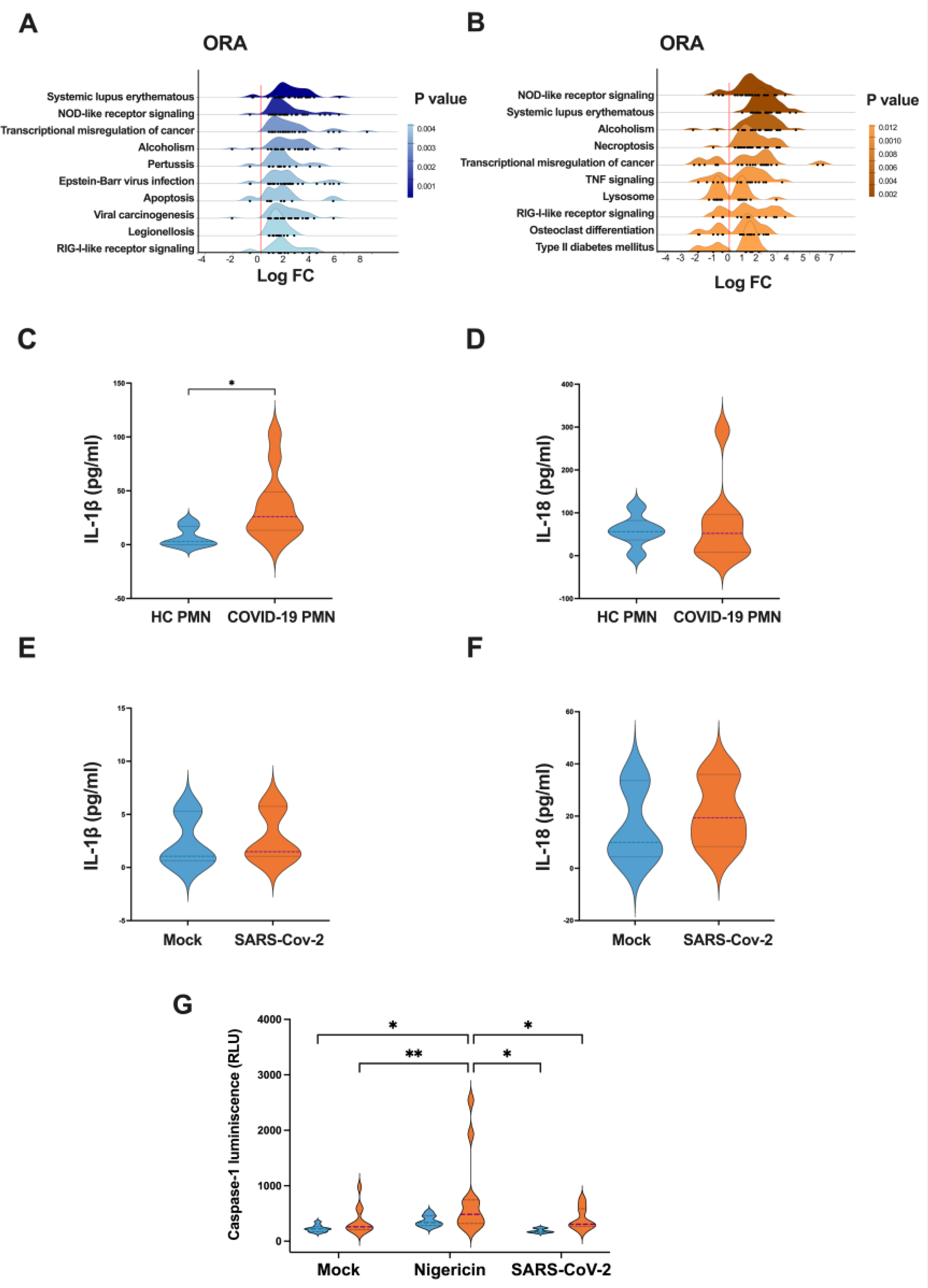
Inflammasome related gene expression and cytokine secretion in PMNs during severe COVID-19. (**A-B**) Ridgeline diagrams of overrepresentation analyses (ORA) according to KEGG database, depicting the top 10 enriched signaling pathways in PMNs during severe COVID-19 compared to (**A**) healthy controls and (**B**) mild COVID-19. (**C**) IL-1β and (**D**) IL-18 levels in 24-h cell culture supernatants from COVID-19 (n = 11 for IL-1β and 9 for IL-18) and HC PMNs (n =6 for both). (**E**) IL-1β and (**F**) IL-18 levels in 24-h cell culture supernatant from PMNs exposed or non-exposed to purified SARS-CoV-2 viral particles (10 virus particles / PMN) (n = 3). (**G**) Caspase1 activity in PMNs following a 2-h stimulation with nigericin or purified SARS-CoV-2 viral particles (10 virus particles / PMN). For HC PMNs, n = 9 for mock and nigericin and n = 6 for SARS-CoV-2 exposure. For COVID-19 PMNs, n = 12 for mock and nigericin and n = 9 for SARS-CoV-2 exposure. *p < 0.05 and **p < 0.01. Data presented as mean ± SD. Tukey’s multiple comparisons test for mixed-effect analysis was applied for (G), meanwhile P values for (C-F) were calculated with the Mann-Whitney U-test.

Given the strong upregulation of many inflammasome related genes during severe COVID-19, we assessed whether PMNs exhibit active inflammasome formation *in vivo*. To evaluate spontaneous inflammasome mediated cytokine secretion, fresh PMNs isolated from severe COVID-19 patients and HC were cultured *ex vivo* overnight. We measured the levels of IL-1β and IL-18 in the supernatant and found that IL-1β secretion was significantly increased in the supernatant of severe COVID-19 PMNs compared to HC PMNs (Figure 2C), whereas the IL-18 levels did not differ significantly (Figure 2D). Additionally, since SARS-CoV-2 viral particles were previously implicated to induce inflammasome formation in macrophages (17), the IL-1β and IL-18 levels after HC PMNs exposure to SARS-CoV-2 were also assessed but no significant effects in the secretion of these cytokines were observed (Figure 2E, F).

The spontaneous secretion of IL-1β by COVID-19 PMNs suggests that these cells are actively producing and releasing IL-1β through inflammasome formation which is dependent on caspase1 activity (32). We assessed caspase1 activity in response to the second signal required for inflammasome activation, induced by nigericin, and observed increased caspase1 activity in severe COVID-19 PMNs compared to HC PMNs (Figure 2G). These findings suggest that severe COVID-19 PMNs have an increased capacity for inflammasome activation, potentially due to an existing priming signal during acute disease *in vivo*. However, no significant difference in caspase1 activity between non-exposed and virus-exposed PMNs were observed (Figure 2G), indicating that caspase1 activation in COVID-19 PMNs is not directly triggered by the virus.

### Activation of neutrophil inflammasome related pathways during respiratory distress is not specific to COVID-19

We also reanalyzed the RNA-seq data generated by LaSalle *et al.* (8), focusing on neutrophil transcriptomics in patients with COVID-19 as compared to non-COVID-19 patients, and healthy controls. The non-COVID-19 patients were presented with acute respiratory distress and clinical concern for COVID-19 but tested negative for SARS-CoV-2 by PCR. Our analysis included IFN-α response, IL-1β production, TLR signaling, NLRP3 inflammasome, and pyroptosis pathways, using the Gene Ontology (GO) database; the NLR signaling pathway using the Kyoto Encyclopedia of Genes and Genomes (KEGG) database; and inflammasome pathway using the REACTOME database (Figure 3). These pathways were significantly enriched in COVID-19 patients, supporting our findings. Importantly, the genes from the above-mentioned pathways were also induced in non-COVID-19 patients, suggesting that these pathways represent a general neutrophil response to inflammatory stimuli rather than a COVID-19 specific response.

**Figure 3.**
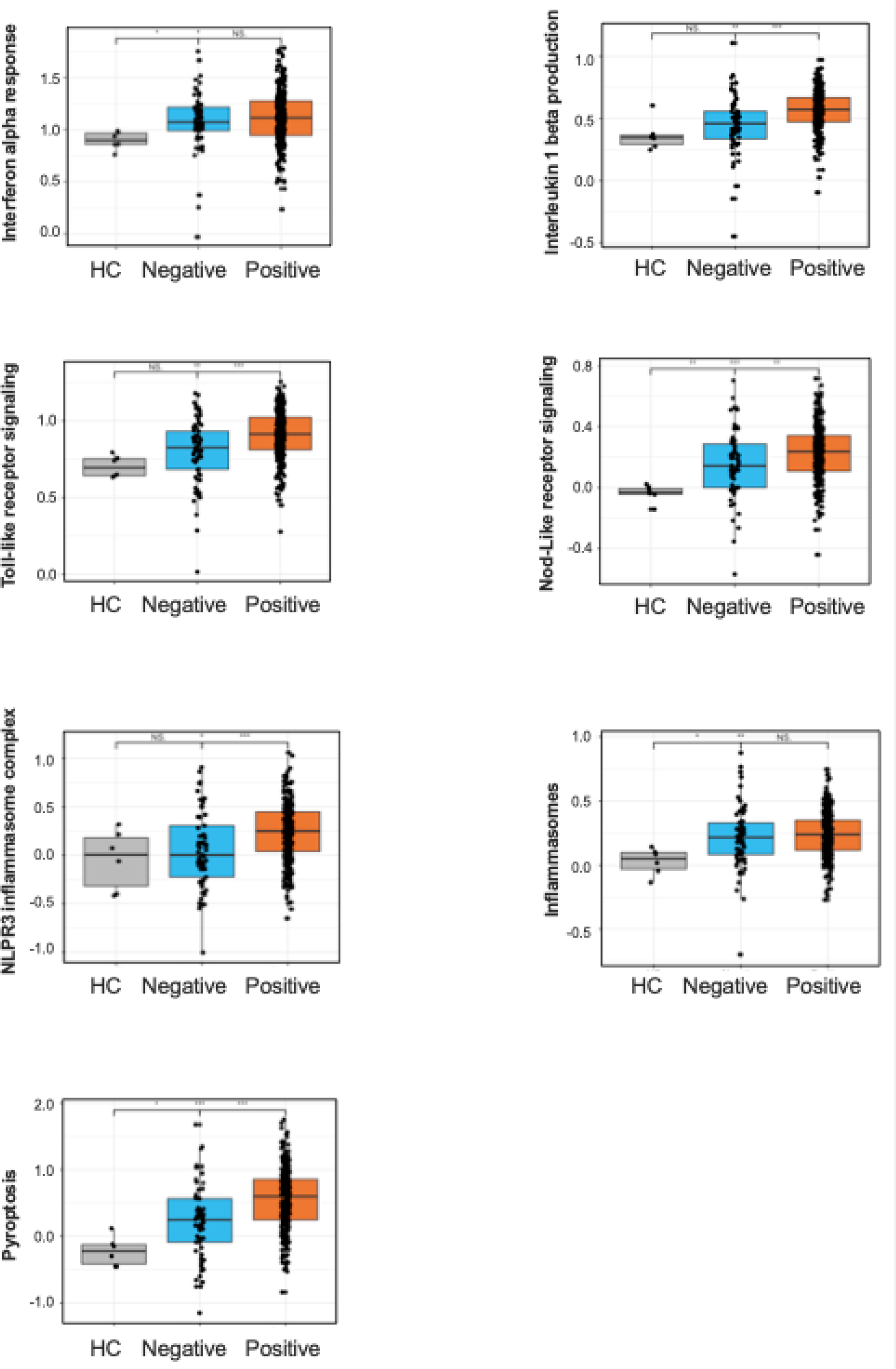
Comparative neutrophil transcriptomics of COVID-19 and non-COVID-19 patients. Bar graphs represent the activation levels of selected pathways and processes as identified by neutrophil transcriptomics. The analysis includes IFN-α responses, IL-1β production, TLR signaling, NLRP3 inflammasomes, and pyroptosis as determined through the Gene Ontology (GO) database. The NOD-like receptor signaling pathway was investigated using the Kyoto Encyclopedia of Genes and Genomes (KEGG) database, and the inflammasome pathway was explored via the REACTOME database. The graphs compare the activation levels of these pathways in healthy controls (HC), non-COVID patients with similar symptoms (COVID-19 negative), and COVID-19 positive individuals. Statistical significance is denoted as follows: *p < 0.05, **p < 0.01, ***p < 0.001, ****p < 0.0001. P values were calculated with Kruskall-Wallis test.

### Type I IFNs prime PMNs for inflammasome activation

Since PMNs from COVID-19 patients concomitantly display a strong IFN-I signature (Figure 1B-C) and an increased propensity for inflammasome activation, we hypothesized that IFN-I could act as the priming signal for PMN inflammasomes during COVID-19. Isolated HC PMNs were stimulated *ex vivo* with exogenous IFN-I and the well-described inflammasome priming (1^st^ signal) and activator (2^nd^ signal) agents LPS and nigericin, respectively (33, 34). After stimulation, both priming signals induced pro-IL-1β (31 kDa) in the cell lysates, followed by the release of active IL-1β (17 kDa) into the supernatant in response to nigericin (Figure 4A), confirming the ability of IFN-I to prime PMNs for inflammasome activation.

**Figure 4.**
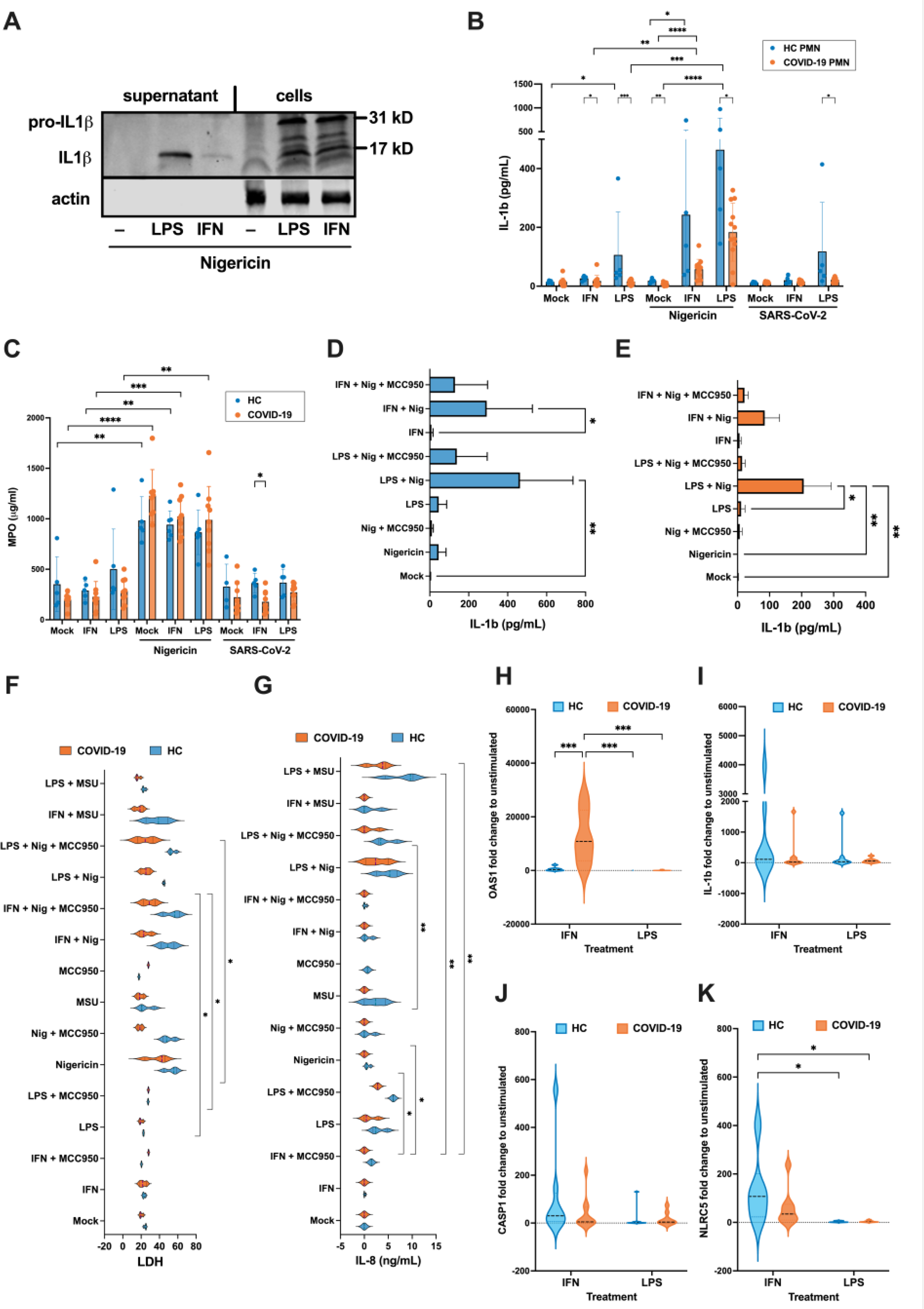
IFN-I primes inflammasome activation while COVID-19 PMNs show defective inflammasome responses *ex vivo*. Isolated HC or COVID-19 PMNs were non-stimulated or stimulated 4h with IFN-I (combination of 2.7*10^4^ IU/ml IFN-α and IFN-β) or 20 ng/ml LPS (1^st^ signal), followed by 4h with 2.5 µM nigericin or purified SARS-CoV-2 (10:1 virus/PMNs) (2^nd^ signal). Then, (**A**) western blot of pro-IL-1β (31 kD) and active IL-1β (17 kD) was performed from HC PMNs supernatant and cell lysates, (**B**) IL-1β (n = 5 HC PMN and 9 COVID-19 PMN) and (**C**) MPO (n= 5 HC PMN and 9 COVID-19 PMN) were measured from supernatants by ELISA. (**D-E**) Effect of inflammasome inhibitor MCC950 (2 µg/ml, added simultaneously with nigericin) on IL-1β secretion in HC (**D**) and severe COVID-19 PMN (**E**) supernatant (n = 3). (**F**) LDH and (**G**) IL-8 in HC and severe COVID-19 PMN supernatants (n = 3). (**H-K**) RT-qPCR of selected mRNAs in IFN-I or LPS-primed HC and COVID-19 PMNs (n = 6-8 HC PMN and 7-10 COVID-19 PMN). *p < 0.05, **p < 0.01, ***p < 0.001, **** p < 0.0001; n = 2-4. Data were presented as mean ± SD. P values calculated with Kruskall-Wallis test for the comparison between treatments by group (HC or COVID-19 PMNs), and Mann-Whitney test for the comparison between HC and COVID-19 PMNs by individual treatment for (B-G), and Two-way ANOVA Tukey’s multiple comparisons test for (B, H-K). Data presented as mean ± SD.

To assess inflammasome formation in circulating neutrophils during COVID-19, PMNs from HC and COVID-19 patients underwent similar stimulation assays as above, followed by IL-1β measurement from supernatants by ELISA. In addition, to further assess the role of SARS-CoV-2 virus particles in neutrophil inflammasome activation, HC PMNs were cultured in the presence of purified viruses (10 infectious units/PMN). HC PMNs responded to both LPS and IFN-I by increasing their IL-1β secretion, which was exponentiated after exposure to nigericin (Figure 4A-B), confirming the ability of IFN-Is to prime for inflammasome assembly in PMNs, albeit less efficiently than LPS. Interestingly, COVID-19 PMNs produced less IL-1β than HC PMNs upon exogenous inflammasome activation primed by either LPS or IFN-I, while SARS-CoV-2 particles did not have any effect on PMN inflammasome activation (Figure 4B). As with 24-h cultures (Figure 2D), we did not detect any significant changes in IL-18 secretion in either HC or COVID-19 PMNs (Supplementary Fig. 4A). However, the release of myeloperoxidase (MPO), used as a marker of degranulation and/or NETosis, in response to nigericin was similar between COVID-19 PMNs and HC PMNs, and therefore the observed diminished IL-1β release by COVID-19 PMNs is not due to general cellular inertia but may be specific to the *ex vivo* induced inflammasome pathway. Furthermore, additional stimulation assays in the presence of the NLRP3 inhibitor MCC950 (Figures 4D-E) and caspase1 inhibitor YVAD (Supplementary Fig. 4B) confirmed that induced IL-1β secretion is dependent on canonical NLRP3 inflammasome activation. Unlike IL-1β (Supplementary Fig. 4C), increased IL-18 secretion was not detectable even after 24-h stimulation (Supplementary Fig. 4D). Furthermore, the observed residual IL-18 was not affected by inflammasome inhibitors, suggesting its secretion to be unrelated to inflammasome activity in PMNs.

We further assessed the specificity of inflammasome activation by measuring LDH and IL-8 levels in the supernatants from the same cells and under the same experimental conditions as shown in Figures 4D-E. The measurements of the former were done to assess inflammasome mediated cell death by pyroptosis in response to nigericin, while the latter was assessed to demonstrate the responsiveness of PMNs to an inflammasome unrelated inflammatory cascade. As with IL-1β secretion, COVID-19 PMNs were less responsive than HC PMNs to nigericin- and LPS-mediated LDH (Figure 4F) and IL-8 (Figure 4G) release, respectively. This suggests that COVID-19 PMNs are generally poorly responsive to inflammatory stimuli.

To examine this reduced responsiveness to external inflammatory priming, we evaluated the inflammasome related gene expression following *ex vivo* stimulation with IFN-I or LPS (Figure 4H-K and Supplementary Fig. 4E-H). OAS1 gene, an interferon stimulated gene (ISG), showed significant upregulation by IFN-I in COVID-19 PMNs as compared to HC PMNs (Figure 4H), while the inflammasome related genes IL-1β (Figure 4I), CASP1 (Figure 4J) and NLRC5 (Figure 4K) were more efficiently induced in HC PMNs than COVID-19 PMNs. This suggests that the inflammasome defect in COVID-19 PMNs is at the transcriptional level when using IFN-I as the priming factor, while high OAS1 gene expression indicates transcriptional defect is restricted to individual genes.

### Association between *ex vivo* inflammasome activation and disease severity

Our analysis of the association between *ex vivo* inflammasome activation (caspase1 activity and IL-1β release) and clinical markers of disease severity, including neutrophil responses, revealed intriguing links. Calprotectin is a marker of neutrophil activation or death (35) but also potentially activates the inflammasome (36). A significant positive correlation between calprotectin plasma levels and PMN caspase1 activity (Figure 5A-B) underscores this latter possibility and highlights the interplay between inflammation and inflammasome activation in PMNs of COVID-19 patients. Furthermore, the negative association of PMN IL-1β levels (after *ex vivo* stimulation with IFN and nigericin) with disease severity (WHO ordinal scale, Figure 5A) and patient neutrophil counts (Figure 5A, C) supports the exhaustion hypothesis, wherein PMNs from severe COVID-19 patients may be less responsive to stimuli due to prior *in vivo* activation. While these findings provide intriguing insights into the complex interplay between calprotectin release, caspase1 activity, and inflammasome activation in COVID-19, additional research is required to further elucidate these connections.

**Figure 5.**
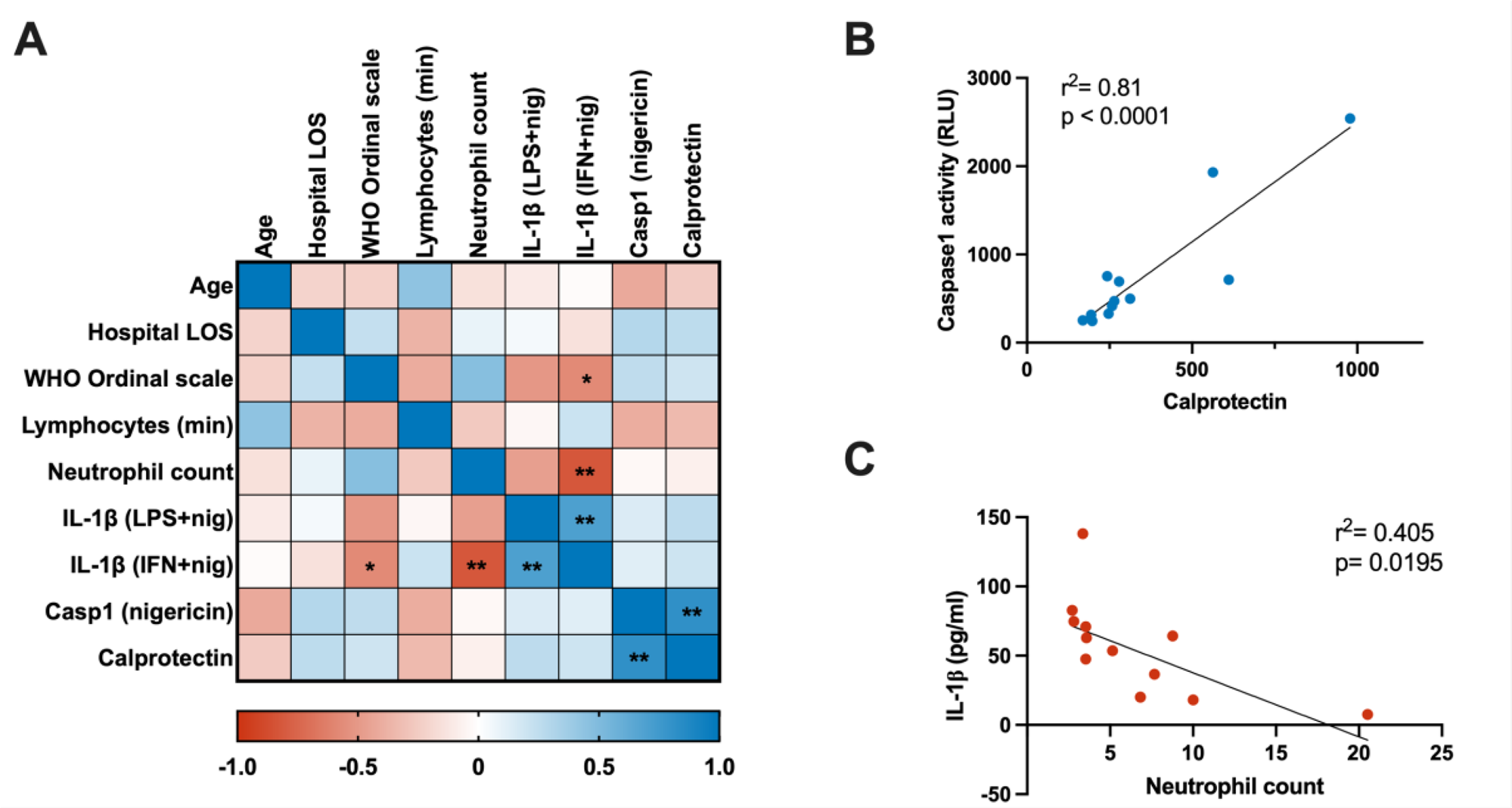
Correlation analysis between clinical parameters and *ex vivo* PMN inflammasome activation. (**A**) Spearman’s correlation matrix depicting the relationships among clinical parameters and results of *ex vivo* experimentation. For the WHO ordinal scale, the baseline parameters were used. (**B-C**) Linear regression analysis demonstrating the associations between: (**B**) Positive association between PMN Caspase1 activity, measured after *ex vivo* nigericin stimulation, and the levels of Calprotectin in the matched patient’s peripheral blood; (**C**) Negative association between e*x vivo* stimulated PMN IL-1β levels (LPS+Nig) and the blood neutrophil count in matched patients at the time of sampling (n=12). *LOS = length of stay. WHO = World Health Organization. Min = minimum. Casp1 = caspase1. LPS or IFN + nig = lipopolysaccharide or type I interferon + nigericin ex vivo stimulation*.

### LDGs differ from PMNs in gene expression and release of inflammasome related interleukins

To assess the inflammasome related inflammatory profile of COVID-19 LDGs in comparison to PMNs, we analyzed the differential expression of inflammasome related genes using RNA-seq (Figure 6A). LDGs differed significantly from PMNs, with the most striking difference being their increased expression of IL-18 and NLRC4, whereas PMNs displayed higher levels of IL-1β, NLRP3 and caspases 1, 4 and 5. Single cell sequencing data from the COVID-19 immune atlas confirmed our transcriptomic results (Figure 6B), from which a detailed gene by gene analysis of the most relevant inflammasome related genes is shown (Figure 6C and Supplementary Fig. 5). Briefly, PYCARD gene coding for the ASC protein was expressed similarly in mature and immature neutrophils (Supplementary Fig. 5), confirming that both cell types have inflammasome forming capacity. However, most of the inflammasome gene expressions differed significantly and in the same manner as in our transcriptomic analysis.

**Figure 6.**
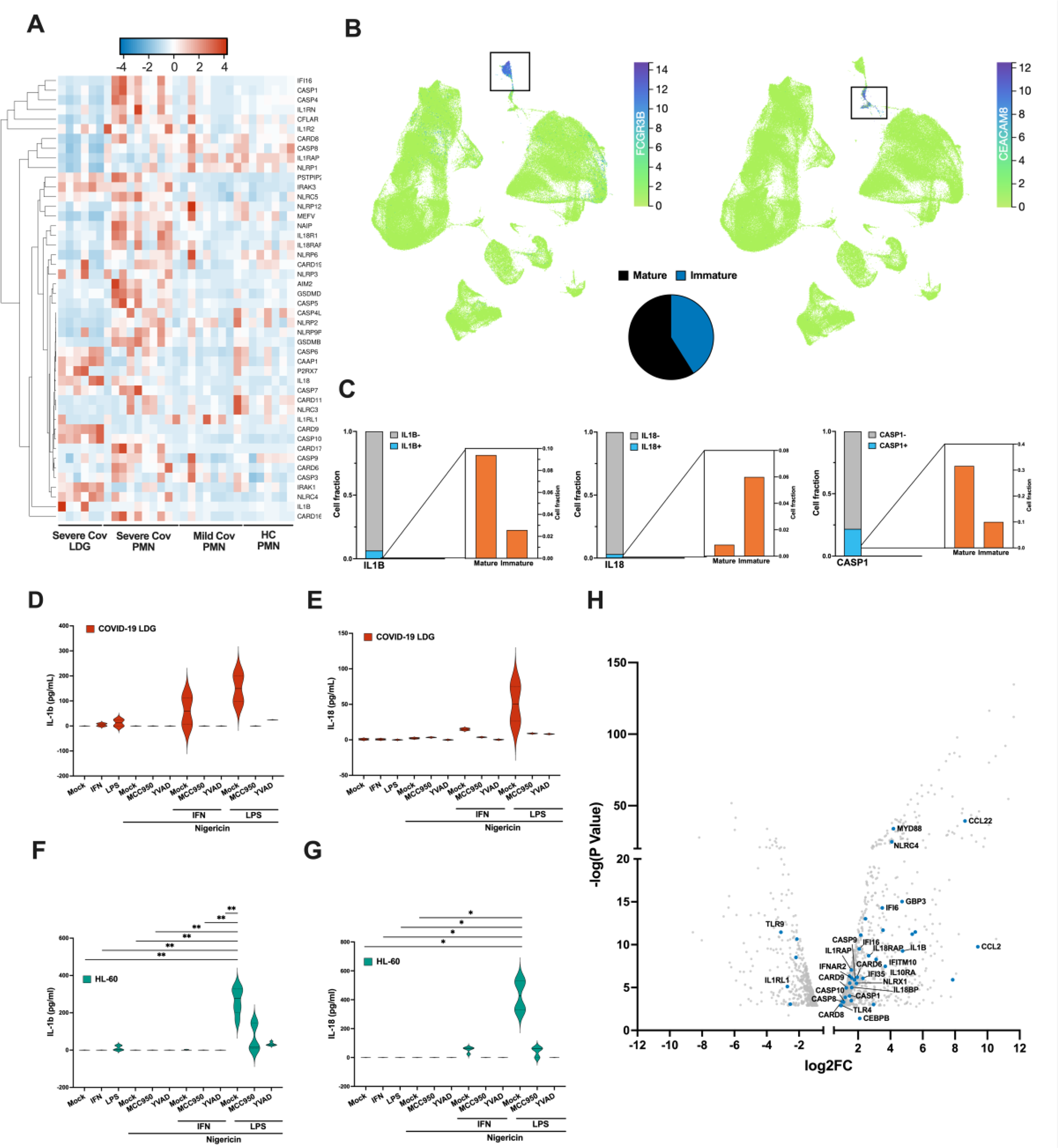
Immature neutrophils express IL-18 in response to inflammasome activation. **(A)** Heatmap depicting selected differentially expressed inflammasome related genes from RNA sequencing performed in PMNs from HC, mild and severe COVID-19, as well as severe COVID-19 LDGs. Only the samples with the highest purity, determined by a cell fraction over 0.65 of neutrophils (identified by CIBERSORTx) are included. The heatmap was clustered by complete linkage and ordered by Spearman’s rank. **(B)** UMAP analysis of the COVID-19 Immune Atlas, which integrates 5 public COVID-19 PBMC single-cell transcriptomics datasets, created using CELLxGENE. (Top) UMAP showing the clustering of CD16^+^ cells (mature, FCGR3B expressing cells) and CD66b^+^ cells (immature, CEACAM8 expressing cells). Each dot represents a single cell colored according to the expression level of a selected gene. The color scale ranges from green (low expression) to purple (high expression). (Bottom) Pie chart summarizing the percentage of mature (black) and immature (blue) cells in the dataset. **(C)** The fraction of mature and immature neutrophils cells expressing inflammasome related genes identified in Figure 7B are shown in a bar graph. For each gene, the proportion of expressing cells is shown in light blue, while the proportion of negative or not-expressing cells is shown in gray. Zoomed-in bar graph depicts the proportion of mature and immature cells expressing each gene. (**D-G**) Isolated COVID-19 LDGs or HL-60 cells (differentiated for 5 days with 1% DMSO) were non-stimulated or stimulated 4h with IFN-I or LPS (1^st^ signal), followed by 4h with nigericin (2^nd^ signal) in the presence or absence of inflammasome inhibitors MCC950 or YVAD as previously. Secretion of (D, F) IL-1β and (E, G) IL-18 were measured from the supernatants by ELISA (n = 2 for LDGs and 3-5 for HL-60). *p < 0.05 and **p < 0.01. P values calculated with Kruskal-Wallis test. Data presented as mean ± SD. (**H**) Volcano plot of differentiated vs undifferentiated HL-60 cells gene expression from GSE93996, with inflammasome related genes marked in blue. Only significant DE genes are shown (adjusted p value < 0.05).

We conducted *ex vivo* stimulation assays using LDGs isolated from COVID-19 patients, similar to the approach used for PMNs described earlier. Like PMNs, IL-1β secretion by LDGs was elevated in the presence of a priming signal (IFN-I or LPS), which exponentially increased when the inflammasome activation signaling molecule nigericin was added (Figure 6D). Contrary to PMNs and in line with the transcriptomics data, an increased IL-18 secretion was detected (Figure 6E). Additionally, the secretion of both ILs by LDGs was inhibited in the presence of inflammasome specific inhibitors MCC950 and YVAD (Figure 6F-G).

These findings suggested that the outcome of neutrophil inflammasome activation varies based on cellular maturation state. To explore this further, we conducted *in vitro* stimulation studies using differentiated HL-60 cells, an immature neutrophil-like model (37). Similar to LDGs from COVID-19 patients, HL-60 displayed comparable IL-18 secretion pattern upon LPS or IFN-I stimulation and nigericin-induced activation. Notably, their IL-1β release was only detected with LPS priming (Figures 6F-G). Furthermore, consistent with transcriptomic analysis revealing an upregulation of inflammasome related genes upon differentiation (Figure 6H), the capacity of HL-60 cells to secrete inflammasome related cytokines was differentiation-dependent (data not shown). Overall, these findings suggest that neutrophils may lose the ability to secrete IL-18 during maturation, and release of neutrophil-derived IL-18 occurs primarily in disease states associated with extensive granulopoiesis and increased immature granulocyte counts in the blood, like COVID-19 (38).

### Neutrophils are recruited to the lungs in SARS-CoV-2 infected mice

Hamsters and human ACE2 expressing mice infected with SARS-CoV-2 develop pulmonary inflammation including neutrophil recruitment (39–41). To further assess the role of neutrophils in COVID-19, we utilized a recently developed SARS-CoV-2 mouse model (18). This model employs the MaVie strain, serially passaged in mouse lungs and causing pneumonia similar to human COVID-19 in wild-type BALB-C mice (18). Infected mice started losing weight by day 2 post-infection, with some mice reaching the clinical endpoint of 20% weight loss by day 4 (Supplementary Fig. 7A, includes animals from 3 independent infection experiments, details of animal usage in Table S2). The first experiment (Exp1) was performed to study infection kinetics and Ly-6G+ neutrophil accumulation in lungs. Viral loads were significantly higher at 2 dpi than 4 dpi (Supplementary Fig. 6A), and viral antigen expression, widespread at 2dpi in bronchioles and alveoli, matched this pattern (Supplementary Fig. 6B and D).The extensive viral replication at 2 dpi was associated with degeneration of infected epithelial cells, most prominent in the respiratory epithelium, accompanied by neutrophil (Ly6G+) infiltration (Supplementary Fig. 6D) and a significant increase in the number of neutrophils in the lungs of infected mice compared to PBS-inoculated mice (Supplementary Fig. 6C). Neutrophil numbers significantly decreased by 4dpi but remained higher than controls (Supplementary Fig. 6C). Detailed information on the histological and immunohistochemical features of these mice is provided in Supplementary Table S2. Together, these findings suggest a pivotal role of neutrophils in clearing the virus in SARS-CoV-2 infected mice.

### Neutrophils from SARS-CoV-2 infected mice display IFN-I dependent caspase1 activation

In the second infection experiment (Exp2) we isolated neutrophils from the lungs of infected mice at 2 and 4 dpi, as well as from non-infected mice (inoculated with PBS) for transcriptomic analysis by RNA-seq. PCA showed differences between neutrophils from infected and non-infected mice, with slight variation between the 2 and 4 dpi time points (Figure 7A). These differences were reflected in many DEGs, including several IFN-I responsive and inflammasome related genes, which showed strong upregulation at 2 dpi with slightly lower but still significantly elevated levels at 4 dpi, compared to non-infected mice (highlighted in the DEG heatmap; Figure 7B). The volcano plot (Figure 7C) provided a comprehensive view of the DEG pattern between neutrophils from SARS-CoV-2 infected and mock-infected mice. In addition to confirming the upregulation of IFN-I responsive and inflammasome related genes observed in the heatmap, the plot revealed a broader transcriptional response to viral infection with several additional DEG.

**Figure 7.**
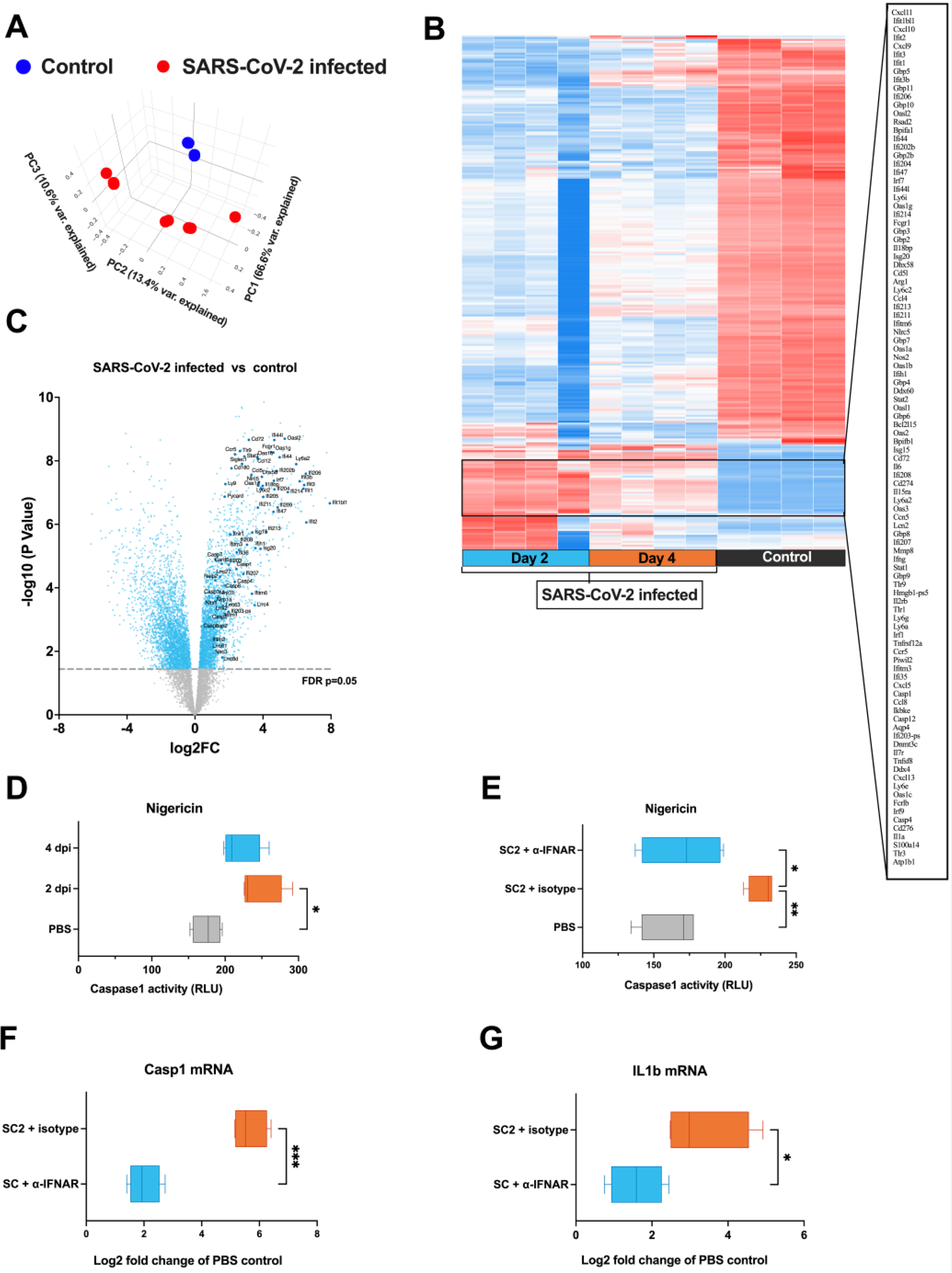
Neutrophils of SARS-CoV-2 infected mice display increased caspase1 activation ability that is dependent on IFN-I. Female BALB/c mice were intranasally inoculated with 5* 10^5^ TCID50 SARS-CoV-2 MaVie strain or PBS as control. Lungs were harvested at 2 and 4 dpi and Ly-6G+ neutrophils isolated based on positive selection with magnetic beads. RNA was isolated and subjected to transcriptomic analysis by RNA-seq. (**A**) Principal component analysis (PCA) of the PBS-inoculated control and SARS-CoV-2 infected mice lung neutrophil RNA-seq samples. (**B**) Heatmap of the top DEGs. (**C**) Volcano plots of DEGs between neutrophils isolated from SARS-CoV-2 infected mice versus uninfected PBS-inoculated mice. Blue points represent significant terms (adjusted p-value < 0.05), while smaller gray points represent non-significant terms. Relevant inflammasome and interferon related genes are shown with larger and darker blue points. **(D)** Caspase1 activity in isolated mice neutrophils following a 2-h stimulation with nigericin was assessed by a bioluminescence method (Caspase-Glo® 1 Inflammasome Assay). (**E-G** Mice were intraperitoneally inoculated with 250 µg anti-IFNAR or IgG1 isotype control directly after infection with SARS-CoV-2 and lung neutrophils isolated at 2 dpi (including also intranasally PBS-inoculated control mice without intraperitoneal injection). **(E)** Caspase1 activity was assessed following a 2-h stimulation with nigericin by bioluminescence method. (**F-G**) RNA was isolated from isolated neutrophils and fold change mRNA expressions of (C) Caspase1 (Casp1) and (D) IL-1β (IL1b) in isotype control and anti-IFNAR treated infected mice as compared to mock-infected control mice assessed by RT-qPCR. DEG = differentially expressed genes. *p < 0.05, and **p < 0.01. P values for D, E and H panels were calculated with ordinary one-way ANOVA using Tukey’s multiple comparisons. Welch’s t-test was used for panels F and G. Data presented as mean ± SD.

We also investigated if neutrophils from SARS-CoV-2 infected mice showed increased inflammasome formation, like PMNs from COVID-19 patients. Neutrophils from infected mice, harvested at 2 and 4 dpi, displayed increased caspase1 activity upon nigericin stimulation, compared to neutrophils from non-infected mice (Exp 2; Figure 7D). The third infection experiment (Exp3) was performed to assess the role of IFN-I by inoculating mice with an IFN-I blocking anti-IFNAR monoclonal antibody or an isotype control antibody post-infection. Remarkably, neutrophils from anti-IFNAR treated mice showed diminished nigericin-induced caspase1 activity (Figure 7E). Furthermore, caspase1 and IL-1β gene expressions were lower in anti-IFNAR treated than isotype treated mice (Figure 7F-G). Taken together, IFN-I appears to be responsible for the increased caspase1 activity in neutrophils of infected mice.

The histology of isotype-treated mice and anti-IFNAR treated mice lungs showed comparable features (Supplementary Fig. 7C) as infections without antibody (Exp 1; Supplementary Fig. 7B-D). Regardless of treatment, some neutrophils in infected mice displayed degeneration and NETosis evidenced by histone H3cit staining (Supplementary Fig. 7C; Supplementary Table S2), and viral loads remained consistent between anti-IFNAR or IgG1 isotype control treated mice (Supplementary Fig. 7B). Taken together, blocking IFN-I signaling did not alter virus replication, virus-induced pathological changes, or early neutrophil recruitment following infection.

## Discussion

Neutrophils, the largest cell population of the host immune system, are rapidly recruited to sites of infection and play an important role in orchestrating an early immune response (42, 43). The relevance of neutrophils in viral infections became increasingly apparent during the COVID-19 pandemic, as they have been shown to be key mediators of the observed pathological processes (44).

This study sheds light on the potential involvement of the inflammasome pathway in COVID-19, particularly by demonstrating its activation in neutrophils during SARS-CoV-2 infection. Our investigation of the inflammatory profile of neutrophils as the dominant population of peripheral blood polymorphonuclear cells (PMNs) revealed an increased ability of neutrophils from severe COVID-19 patients for inflammasome assembly as evidenced by their transcriptional profile, spontaneous release of IL-1β, and elevated caspase1 activity. These findings are consistent with previous reports indicating activation of the NLRP3 inflammasome and ASC specks in circulating neutrophils during acute COVID-19 (14, 16). Furthermore, despite showing increased caspase1 activity, neutrophils from COVID-19 patients exhibited diminished soluble IL-1β production upon exogenous activation of the NLRP3 inflammasome pathway compared to healthy controls, which suggests that this pathway is “exhausted” due to prior activation during the disease. Mechanistically, our findings show that IFN-I, elevated in COVID-19 patients (45, 46), can prime inflammasome formation in neutrophils. Transcriptomic analyses revealed that circulating neutrophils during severe COVID-19 show increased expression of IFN-responsive genes, suggesting inflammasome priming by IFN-I also *in vivo* during COVID-19 (47). Furthermore, the study found that immature neutrophils, which are prevalent in low-density granulocyte fraction (LDGs), exhibit unique inflammasome gene expression and outcomes compared to mature neutrophils (PMNs). LDGs release IL-18 and upregulate distinct inflammasome related genes but lack the IFN-I signature seen in PMNs during COVID-19, indicating lower responsiveness to IFN-I, and supported by less efficient IFN-I mediated inflammasome priming of LDGs *ex vivo*.

SARS-CoV-2 infected mice also showed increased neutrophil caspase1 activity, reversible by an IFN-I receptor (IFNAR) blocking antibody. Transcriptional analysis revealed a robust IFN-I signature and elevated expression of inflammasome genes encoding for caspase1 and IL-1β in neutrophils of infected mice, which were also inhibited by blocking IFNAR signaling, suggesting that IFN-I may also prime for inflammasome activation in mice. Notably, the anti-IFNAR treatment did not affect neutrophil recruitment or NETosis, which is consistent with another COVID-19 model using transgenic human ACE2, where IFNAR knockout inhibited recruitment of monocytes and lymphocytes, but not neutrophils, to infected lungs (48).

Inflammasomes were first studied in macrophages, revealing many molecular mechanisms regulating inflammasome assembly (49). Macrophage inflammasome activation has emerged as a major factor also in COVID-19 (17). Interestingly, macrophage inflammasome activation was recognized to be IFN-I mediated in an experimental rhesus macaque COVID-19 model (50). However, due to the abundance of neutrophils compared with cells of monocyte/macrophage lineage (51, 52), the significance of neutrophil inflammasomes in COVID-19 is likely underestimated. Our results highlight inflammasomes as an additional important inflammatory mechanism in neutrophils (14), complementing their role in phagocytosis, reactive oxygen species generation, degranulation, and NETosis (31).

SARS-CoV-2 can directly activate inflammasomes in cells of the monocyte/macrophage lineage (17). Our study investigated whether SARS-CoV-2 can provide the first or second signal for inflammasome activation in neutrophils. However, we found no evidence of direct virus-induced inflammasome activation in neutrophils. The difference between macrophages and neutrophils in their susceptibility to SARS-CoV-2 could depend on many factors. Both cell types express ACE2, the receptor for SARS-CoV-2, but may differ in ACE2 expression levels (53). Furthermore, the intracellular environment of macrophages is better suited for viral replication (54), while neutrophils focus on phagocytosis and antimicrobial responses (31, 55). Additionally, pathogen opsonization can trigger inflammasomes in macrophages (56) but is not a primary function of neutrophils. Therefore, our findings suggest neutrophil inflammasome activation in response to SARS-CoV-2 likely results from interactions with infected and/or dying cells in the lungs, rather than direct virus activation. To note, whether SARS-CoV-2 can induce neutrophil inflammasomes through immune complex-mediated mechanisms, as seen in monocytes/macrophages (17) remains to be determined.

In this study, we demonstrated IFN-I as the first signal for NLRP3 inflammasome activation in neutrophils. While prior research has explored IFN-inflammasome crosstalk (57), priming capacity of IFN-I remained unclear. While IFN-I promotes inflammasomes in epithelial cells (58) it can also dampen IL-1β in macrophages (59). Plausibly, initial IFN-I exposure may upregulate inflammasome genes, whereas prolonged activity could hinder IFN-I signaling via “negative feedback” loop, in line with our findings of inflammasome exhaustion in circulating neutrophils of severe COVID-19 patients. It should be noted that several SARS-CoV-2 encoded proteins have been shown to inhibit IFN-I signaling (60). However, no evidence suggests that neutrophils can be infected by SARS-CoV-2 and therefore it seems unlikely that such direct virus mediated effects could play a role in the observed neutrophil unresponsiveness to IFN-I.

The dualistic nature of the IFN-I response in COVID-19 has been recognized previously. It seems that a strong initial IFN-I response to SARS-CoV-2 is more likely to result in asymptomatic or mild COVID-19 whereas a decreased initial IFN-I activity, due to e.g. genetic defects or increased levels of IFN-I autoantibodies, can lead to more severe COVID-19 (61).

This initial beneficial effect of IFN-I is probably due to its ability to limit viral replication at early stages of the infection. However, at later stages of the disease IFN-I can be detrimental by promoting inflammatory pathways instead of direct antiviral effects (62). Thus, similarly to the IFN-I response in general, the role of neutrophil inflammasomes in development and severity of COVID-19 might be dualistic in nature with an initial protective effect while damaging when sustained for prolonged periods.

Our study demonstrated a strong association between PMN caspase1 activity and plasma levels of calprotectin, a marker of neutrophil activation. Additionally, increased disease severity, as assessed by the WHO ordinal scale, was significantly linked to PMNs being less responsive to *ex vivo* IFN-induced inflammasome activation. Thus, these results suggest that neutrophil inflammasomes playing a potential role in disease severity rather than being protective in COVID-19.

Our study also unveiled distinct gene profiles in LDGs and PMNs from severe COVID-19 patients. LDGs exhibited upregulation of genes related to DNA replication and cell cycle, indicating immaturity, and confirming our prior findings (7). Conversely, PMNs displayed heightened NLR signaling, suggesting a robust response to pathogens. While our study compared PMNs and LDGs, and the COVID-19 Immune Atlas single cell analysis represented a broader classification of mature and immature neutrophils, the alignment of our results with the atlas provides further support for the distinct characteristics of these two neutrophil populations in severe COVID-19. Notably, IL-18 gene expression and secretion after *ex vivo* stimulation were higher in LDGs than PMNs. To note, PMN’s lack of IL-18 secretion is not due to lack of protein, as they constitutively express significant amounts intracellularly (63). This indicates a similarity between LDGs and monocytes/macrophages in inflammasome mediated IL-18 processing, possibly lost during neutrophil maturation.

The present study has some limitations worth discussing. Firstly, the relatively small human sample size may limit the generalizability of the findings. While RNA-seq provided valuable insights into gene expression profiles of PMNs and LDGs, we did not perform functional validation of the identified pathways in this study. Regarding our experimental SARS-CoV-2 disease model, the high virus input might trigger robust immune responses that differ from typical human infections, and the short-lived virus replication in the applied model does not capture the effect of prolonged antigen exposure or the complex inflammatory milieu seen in human cases. Furthermore, the observed inhibitory effects on neutrophil inflammasome activity by IFNAR blockade does not exclude the possibility that IFN-I could promote neutrophil inflammasome formation by indirect effects such as stimulating the release of pro-inflammatory cytokines by other cell types. Finally, since the prominent role of neutrophils in the immune response to viral infections is widely recognized (42, 43) and it would be valuable to compare these findings to neutrophil responses in other viral respiratory infections.

Taken together, our findings provide valuable insights into neutrophil involvement in COVID-19 and possibly other viral respiratory infections. However, further research is needed to fully grasp the role of neutrophil inflammasomes in COVID-19 pathogenesis. This increased understanding may facilitate the development of targeted treatment approaches for COVID-19. For example, pharmacologically targeting the inflammasome pathway in neutrophils with novel inhibiting molecules (64), may help mitigate the exaggerated inflammatory response observed in severe cases. The next steps involve validating the pathways and genes identified as potential therapeutic targets and assessing their COVID-19 specificity. Prospectively, these strategies could be extended to address upcoming respiratory virus pandemics, where neutrophils and inflammasomes provide major pathogenic contributions.

## Data Availability

Once published, The data supporting the findings of this study will be made available in public repositories such as GEO for RNAseq data. Also, the data will be available upon reasonable request from the corresponding author, which will be provided in accordance with relevant ethical and privacy regulations to ensure the confidentiality and anonymity of study participants.

## Contributors

L.E.C.: conceptualization, data curation, formal analysis, investigation, software, validation, visualization, writing– original draft; S.T.J.: investigation, writing – review & editing; Sa.M.: investigation; Si.M.: investigation; R.K.: resources; L.K.; investigation, writing – review & editing; Ta.S.: resources; J-P.P.: resources; Anu.K.: funding acquisition, resources, writing – review & editing; E.K.: supervision, writing – review & editing; H.L.: resources; P.M.: resources, writing – review & editing; Anj.K.: investigation, methodology, supervision, validation, visualization, writing – review & editing; O.V.: funding acquisition, resources; To.S.: conceptualization, data curation, formal analysis, funding acquisition, investigation, methodology, project administration, supervision, validation, writing – original draft.

## Acknowledgements

This work was financed by grants by the Academy of Finland to To.S. (321809), Anu.K. (336439 and 335527); grants by the Helsinki University Hospital funds to O.V. (TYH 2021343); EU Horizon 2020 programme VEO (874735) to O.V.; Finnish governmental subsidy for Health Science Research (TYH 2021315) to Anu.K.; Paulon Säätiö to L.E.C.; Suomen Lääketieteen Säätiö to L.E.C.; Jane and Aatos Erkko foundation to O.V. The funders had no role in study design, data collection and analysis, nor decision to publish, or preparation of the manuscript.

RNA isolation, library preparations and RNA sequencing was performed at the Institute for Molecular Medicine Finland FIMM, Genomics unit supported by HiLIFE and Biocenter Finland. The authors also thank M. Utriainen for expert technical assistance.

## Supplementary figure legends

**Supplementary Figure 1.**
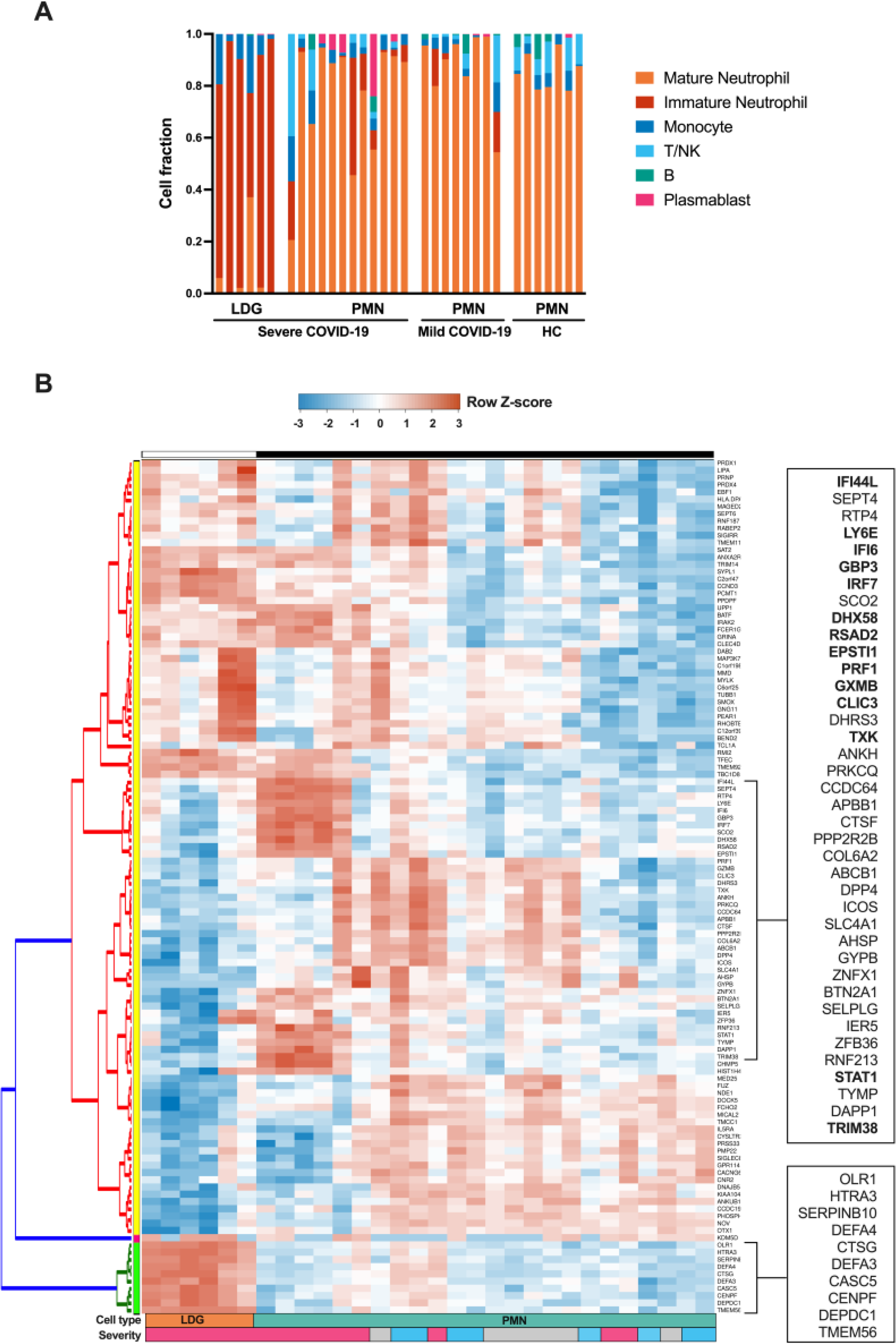
Comparison of gene expression in granulocyte populations of COVID-19 patients using RNA-seq analysis. **(A)** Deconvoluted RNA-seq data. The cellular composition in isolated PMN and LDG fractions was estimated using CIBERSORTx through the identification of cell populations based on RNA-seq. The bar plots in the figure represent the cell composition of each RNA-seq sample, offering insights on sample purity. **(B)** Heatmap of the top 118 differentially expressed genes between PMNs from healthy controls, mild and severe COVID-19, as well as LDGs from severe disease, identified by unsupervised ICGS analysis based on correlation, using AltAnalyze software. IFN-related genes, identified by GENESHOT, are shown in bold.

**Supplementary Figure 2.**
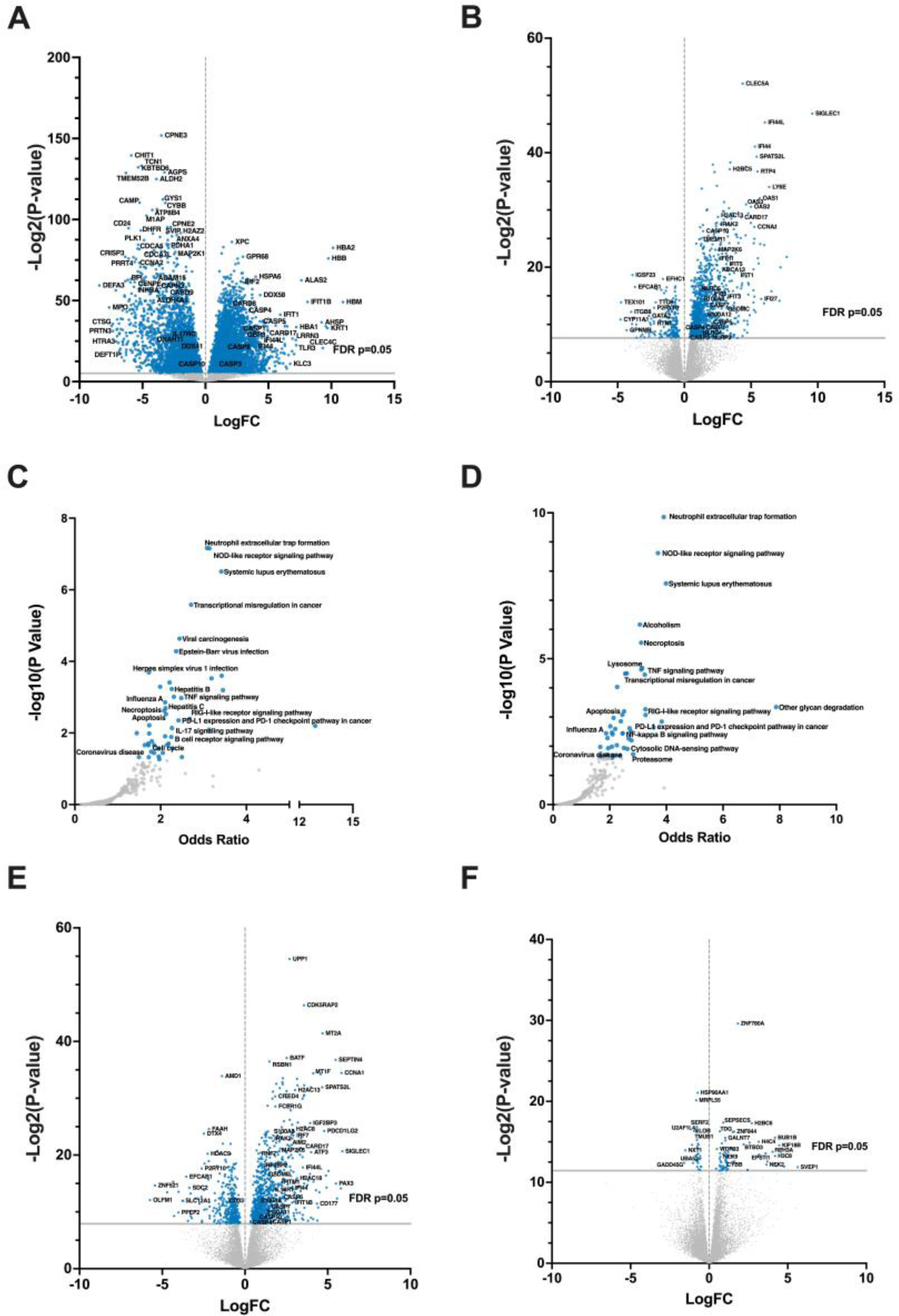
Enriched differentially expressed genes and pathways in severe COVID-19 PMNs and LDGs. (**A-B**) Volcano plots of DEGs between severe COVID-19 PMNs versus (**C**) HC PMNs and (**D**) severe COVID-19 LDGs. (**C-D**) Volcano plots of enriched gene sets in severe COVID-19 PMNs versus (**A**) HC PMNs and (**B**) severe COVID-19 LDGs, using KEGG database. Each point represents a single gene set, where the x-axis measures its odds ratio, while the y-axis shows its -log10(p-value). (**E-F**) Volcano plots of (**E**) severe COVID-19 PMNs versus mild COVID-19 PMNs and (**F**) mild COVID-19 PMNs vs HC PMN. For all panels, blue points represent significant terms (adjusted p-value < 0.05), while smaller gray points represent non-significant terms. *DEG = differentially expressed genes*.

**Supplementary Figure 3.**
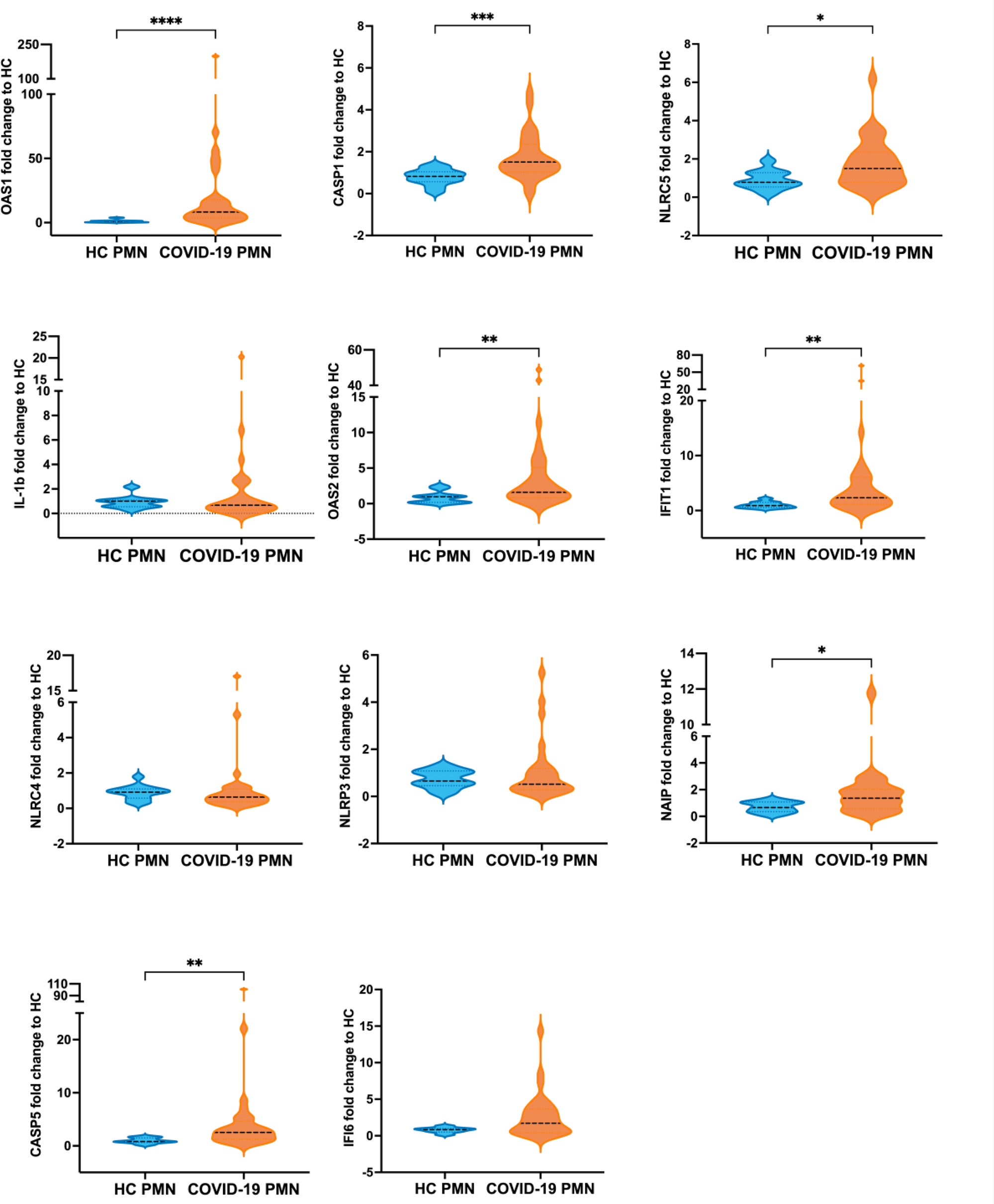
Differential expression of interferon and inflammasome related genes in PMNs during COVID-19. RNA was extracted from isolated HC PMNs (n = 8-13) versus severe COVID-19 PMNs (n = 29-32) and subjected to comparative RT-qPCR using specific primers for OAS1, OAS2, IFIT1, IFI16, caspase1, caspase5, IL1B, NLRC4, NLRC5, NLRP3 and NAIP. *p < 0.05, **p < 0.01, ***p < 0.001 and **** p < 0.0001. P values calculated with Mann-Whitney U-test. Data presented as mean ± SD.

**Supplementary Figure 4.**
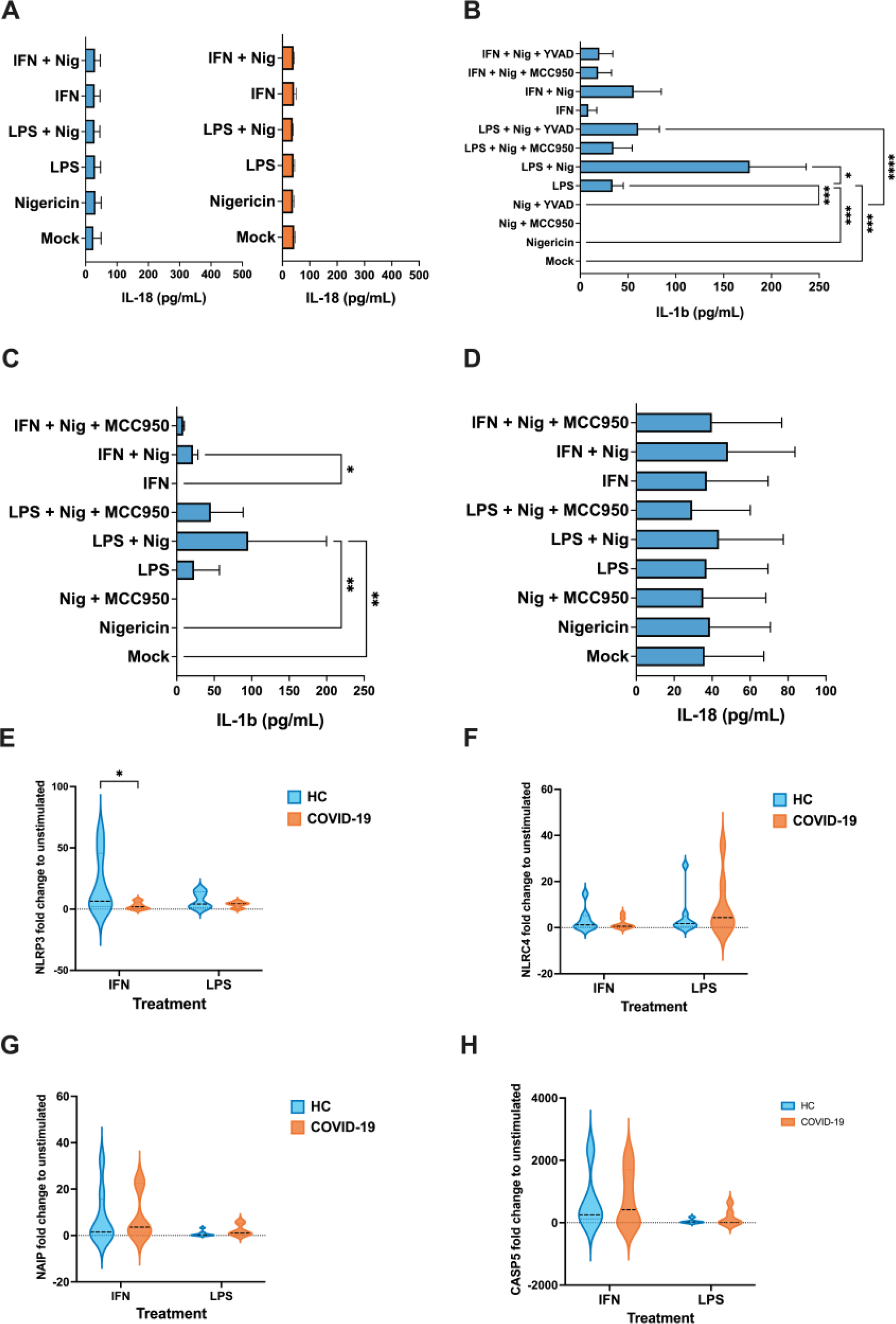
*Ex vivo* stimulation of isolated PMNs. **(A)** IL-18 (n = 2-3 HC PMN and 3 COVID-19 PMN) was measured from supernatants by ELISA following LPS or IFN-I priming (4-h) and subsequent nigericin activation (4-h). (**B-C**) Effect of different inflammasome specific inhibitors in cytokine secretion. (**A**) Effect of inflammasome inhibitor MCC950 (2 µg/ml) and YVAD (20 µg/ml) on LPS or IFN-I primed (4-h) and nigericin activated (4-h) IL-1β secretion in the supernatant of healthy control PMNs (n = 8). (**B-C**) Effect of inflammasome inhibitor MCC950 (2 µg/ml, added simultaneously with nigericin) on LPS or IFN-I primed (4-h) and nigericin activated (20-h) (B) IL-1β and (C) IL-18 secretion in the supernatant of healthy control PMN (n = 3). (**D**) Gene expressions in HC and COVID-19 PMNs after LPS or IFN-I stimulation. A comparison of gene expression in isolated healthy control PMNs versus COVID-19 PMNs after *ex vivo* stimulation with LPS or IFN-I. Extracted RNA was subjected to comparative RT-qPCR using specific primers for NLRP3, NLRC4, NAIP and CASP5 (n = 4-8 for HC PMN and 6-9 for COVID-19 PMN). *p < 0.05. Two-way ANOVA with Tukey’s multiple comparison test was applied. Data were presented as mean ± SD. *p < 0.05, **p < 0.01, ***p < 0.001, **** p < 0.0001. P values calculated with Kruskal-Wallis test. Data presented as mean ± SD. *IFN = interferon type I, LPS = lipopolysaccharide, Nig = nigericin, YVAD = tetrapeptide caspase1 inhibitor Tyr-Val-Ala-Asp*.

**Supplementary Figure 5.**
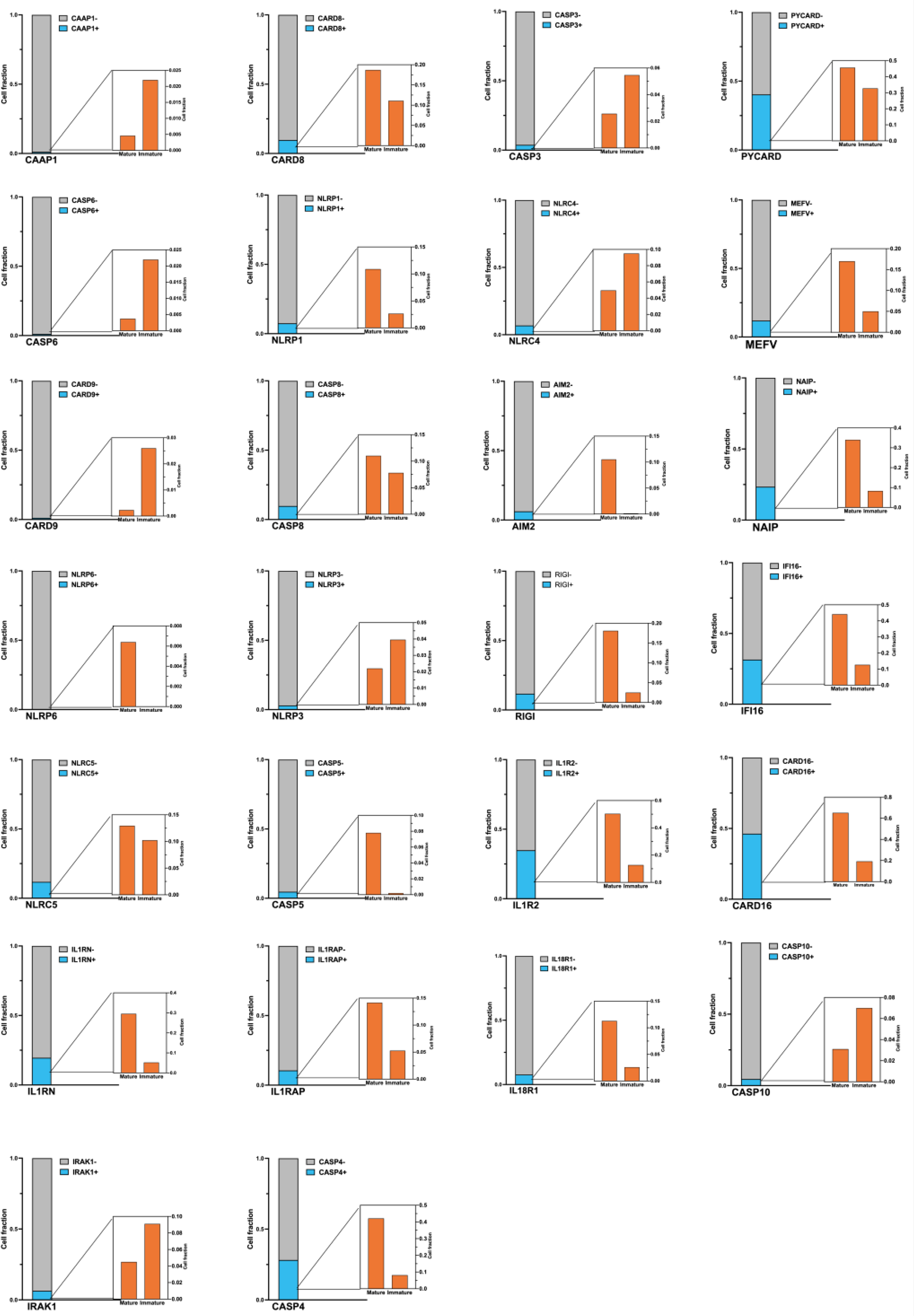
Expression of inflammasome related genes in mature and immature neutrophils from COVID-19 PBMCs. The fraction of mature and immature neutrophils cells expressing 17 inflammasome related genes identified in Figure 7B (shown in black and blue, respectively) are shown in a bar graph. For each gene, the proportion of expressing cells is shown in light blue, while the proportion of negative or not-expressing cells is shown in gray. Zoomed-in bar graph depicts the proportion of mature and immature cells expressing each gene.

**Supplementary Figure 6.**
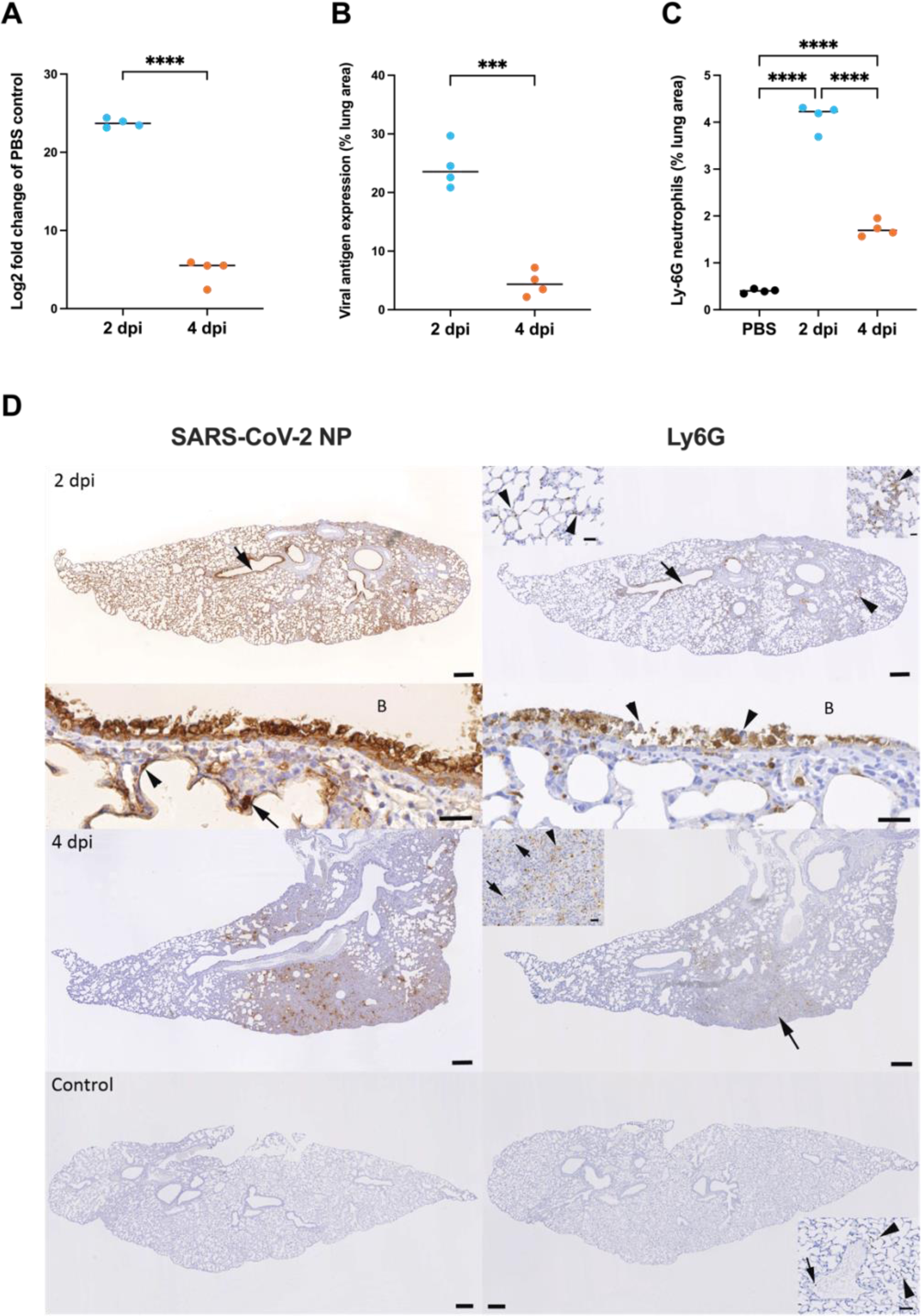
Neutrophil accumulation in the lungs correlates with viral loads in SARS-CoV-2 infected mice. Female BALB/c mice were intranasally inoculated with 5*10^5^ TCID50 SARS-CoV-2 MaVie strain or PBS as control and euthanized at 2 dpi or 4 dpi. **(A)** RNA was isolated from lungs and subjected to RT-qPCR targeting viral subE and GAPDH as housekeeping gene. The relative expression of subE was measured using the comparative Ct method as compared to mock-infected control (in which subE was undetectable but set to 40 Ct) **** p < 0.0001. P values calculated with Welch’s t-test. **(B)** Quantification based on morphometric analysis that determines the area of immunolabelling for SARS-CoV-2 nucleoprotein in relation to total tissue area. ***p < 0.001. P values calculated with Welch’s t-test. Black line represents the mean. **(C)** Quantification of Ly-6G based on morphometric analysis that determines the area of immunolabelling for Ly6G in relation to total tissue area in mock-infected controls. **** p < 0.0001. P values calculated with ordinary one-way ANOVA using Tukey’s multiple comparisons test. Black line represents the mean. **(D)** Left column: immunohistochemistry for SARS-CoV-2 nucleoprotein; right column: immunohistochemistry for Ly6G (neutrophil marker), hematoxylin counterstain. Bars = 500 µm (large images) and 50 µm (insets). At 2 dpi (top), the arrow points at a bronchus with viral antigen expression in epithelial cells. A close–up of the bronchus (bottom; B: bronchial lumen) shows degenerated and slough off antigen positive epithelial cells. Adjacent alveoli exhibit viral antigen expression in typeI (arrowhead) and typeII (arrow) pneumocytes. The overview (top) shows neutrophils between the infected bronchial (arrow) epithelial cells, in parenchymal areas (arrowhead; right inset) and in capillaries (arrowheads). A close-up of the bronchus (bottom; B: bronchial lumen) highlights numerous neutrophils between degenerate (arrowheads) epithelial cells. At 4 dpi (middle), there are focal areas with antigen expression in alveolar epithelial cells and infiltrating macrophages. Neutrophils are present among the infiltrating cells (arrow) as individual cells (inset: arrows) or in aggregates (inset: arrowhead). The bottom shows the lung of a mock-infected control animal. There is no viral antigen expression. Staining for Ly6G depicts individual neutrophils in larger vessels (inset: arrow) or in capillaries (inset: arrowheads).

**Supplementary Figure 7.**
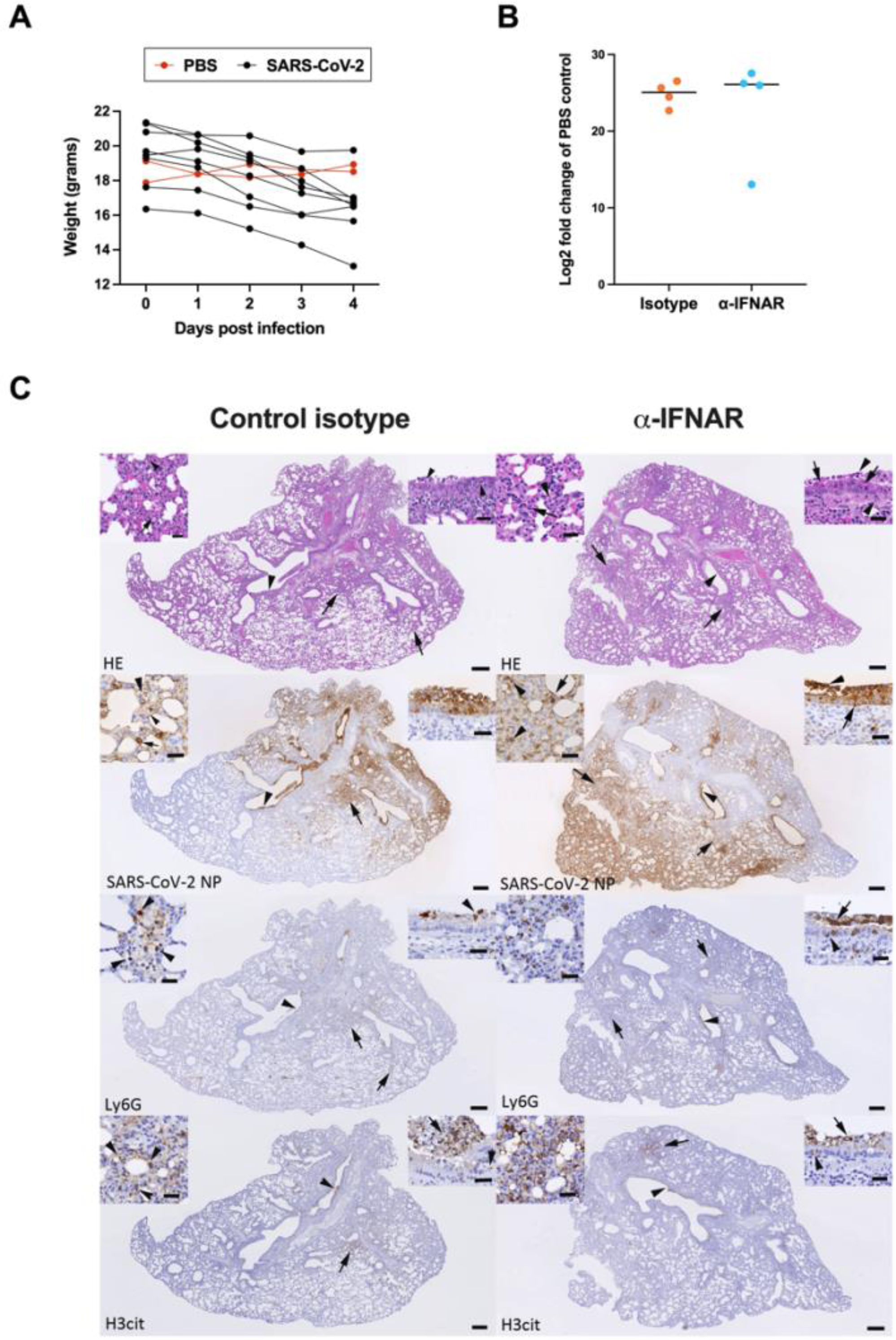
Dynamics of animal weight and impact of α-IFNAR treatment in SARS-CoV-2 infected mice. Female BALB/c mice were intranasally inoculated with 5* 10^5^ TCID50 SARS-CoV-2 MaVie strain or PBS as control. **(A)** Daily tracking of animal weight performed throughout the experiment (n = 8 for SARS-CoV-2 infected animals, n = 2 for PBS-inoculated animals). The weights of the mice euthanized at 2 dpi (n= 26) did not show significant differences and are not reported. (**B-C**) Mice were intraperitoneally inoculated with 250 µg anti-IFNAR or IgG1 isotype control directly after infection with SARS-CoV-2 and lung neutrophils isolated at 2 dpi (including also intranasally PBS-inoculated control mice without intraperitoneal injection) **(B)** RNA was isolated from mouse lungs and subjected to RT-qPCR targeting the replication-intermediate subgenomic E gene and GAPDH as housekeeping gene. RNA levels were assessed based on cycle threshold Ct levels. The expression levels of the target gene SubE were measured and normalized to GAPDH levels using the comparative Ct method (ΔΔCt). The fold change values were calculated by the formula 2^(−ΔΔCt), representing the relative gene expression compared to the PBS mock-infected control (in which subE was undetectable but set to 40 Ct). No significant differences are seen between the two groups, assessed with Welch’s t-test. **(C)** Histological features, viral antigen expression and extent of neutrophil influx and damage in the lung of SARS-CoV-2 infected BALB/C mice after isotype control and anti-IFNAR treatment at 2dpi. Left column: Control isotype treated mice; right column: anti-IFNAR treated mice. HE stain (top layer) and immunohistology, hematoxylin counterstain (all other images). Bars: 250 µm (overview images) and 25 µm (insets). In control isotype treated mice, the lung exhibits degeneration and loss of bronchial and bronchiolar epithelial cells (HE stain: arrowhead; right inset), with mild inflammatory infiltration. The parenchyma exhibits focal areas of increased cellularity, with typeII pneumocyte activation and occasional degenerate alveolar epithelial cells (arrows; left inset: degenerate cells (arrowhead) and infiltrating neutrophil (arrow)). Staining for SARS-CoV-2 NP confirms epithelial cell infection in bronchus (arrowhead; right inset) and alveoli (arrow; left inset). Right inset: Viral antigen expression is seen in intact and sloughed off, degenerate epithelial cells. Left inset: Viral antigen expression is seen in both typeI (small arrowhead) and typeII (small arrow) pneumocytes; there are also degenerate positive cells (large arrowhead). Neutrophils (Ly6G+) are located within focal parenchymal areas of increased cellularity (arrows; left inset: arrowheads) and present between degenerate bronchial epithelial cells (arrowhead; right inset: arrowhead). Staining for histone H3 shows neutrophil degeneration/NETosis in parenchymal areas (arrow; left inset: arrowheads) and associated with degenerate epithelial cells (arrowhead; right inset: positive reaction between sloughed off epithelial cells (arrow) and between the intact epithelial layer (arrowhead)). In anti-IFNAR treated animals, the lung exhibits degeneration and loss of bronchial and bronchiolar epithelial cells (arrowhead; right inset: arrows), with mild inflammatory infiltration and individual neutrophils between intact and sloughed off degenerate epithelial cells (right inset: arrowheads). The parenchyma exhibits focal areas of increased cellularity, with typeII pneumocyte activation and occasional degenerate alveolar epithelial cells (arrows; left inset: degenerate cells (arrow) and infiltrating neutrophils (arrowhead). Staining for SARS-CoV-2 NP shows epithelial cell infection in bronchioles (arrowhead; right inset) and alveoli (arrow; left inset). Right inset: Viral antigen expression is seen in intact and sloughed off, degenerate epithelial cells. Left inset: Viral antigen expression is seen in pneumocytes (arrow) and infiltrating macrophages (arrowheads). Neutrophils (Ly6G+) locate within focal parenchymal areas of increased cellularity (arrows; left inset) and are present between intact (inset: arrowhead) and degenerate epithelial cells (arrowhead; right inset: arrow). Staining for histone H3cit shows neutrophil degeneration/NETosis in parenchymal areas (arrow; left inset) and associated with degenerate epithelial cells (arrowhead; right inset: positive reaction between sloughed off epithelial cells (arrow) and between the intact epithelial layer (arrowhead)). *Dpi= days post infection; NP = nucleoprotein*.

## Tables & table legends

**Supplementary Table S1.**
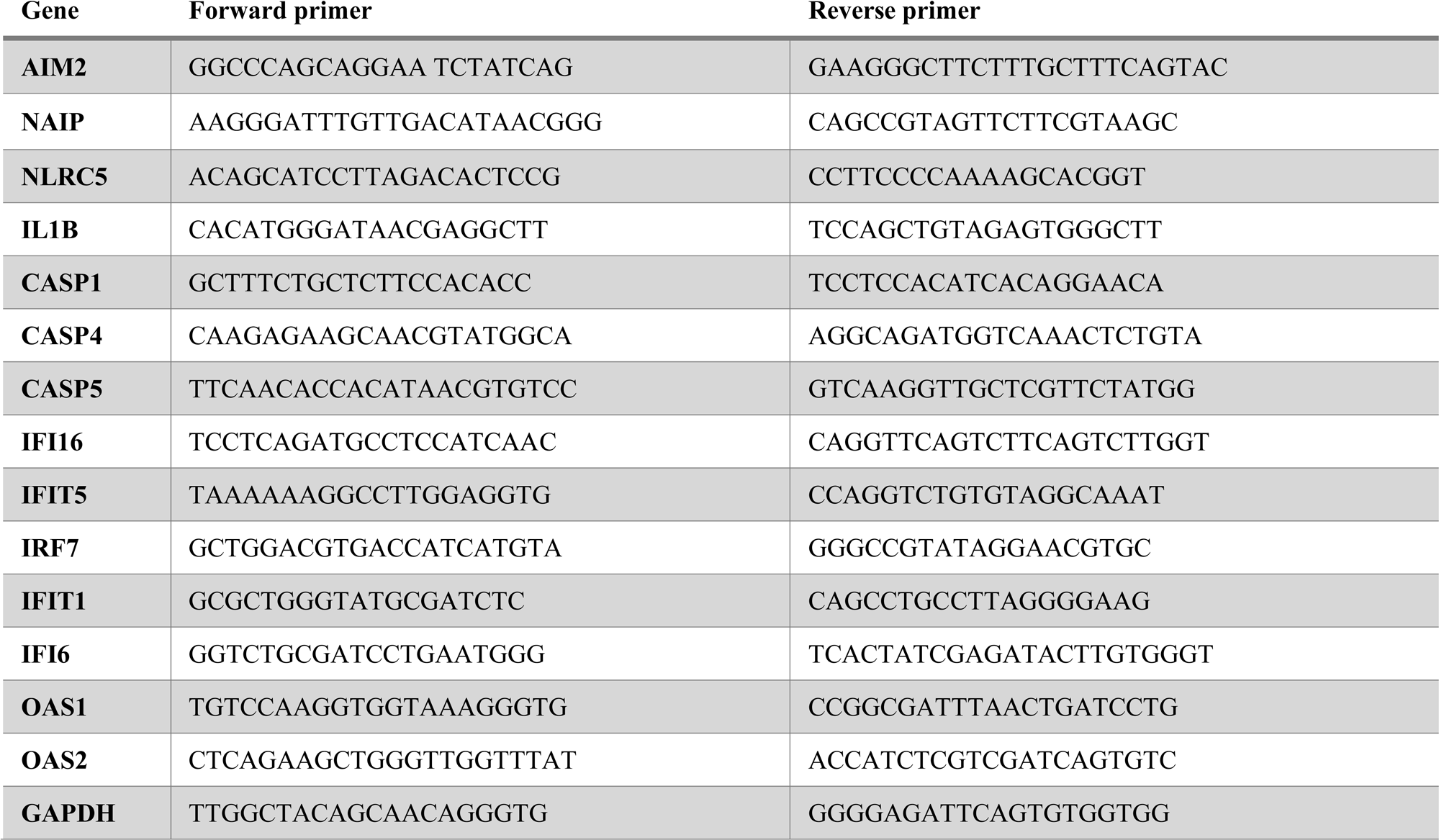
qPCR primer sequences: gene-specific forward and reverse primers.

**Supplementary Table S2.**
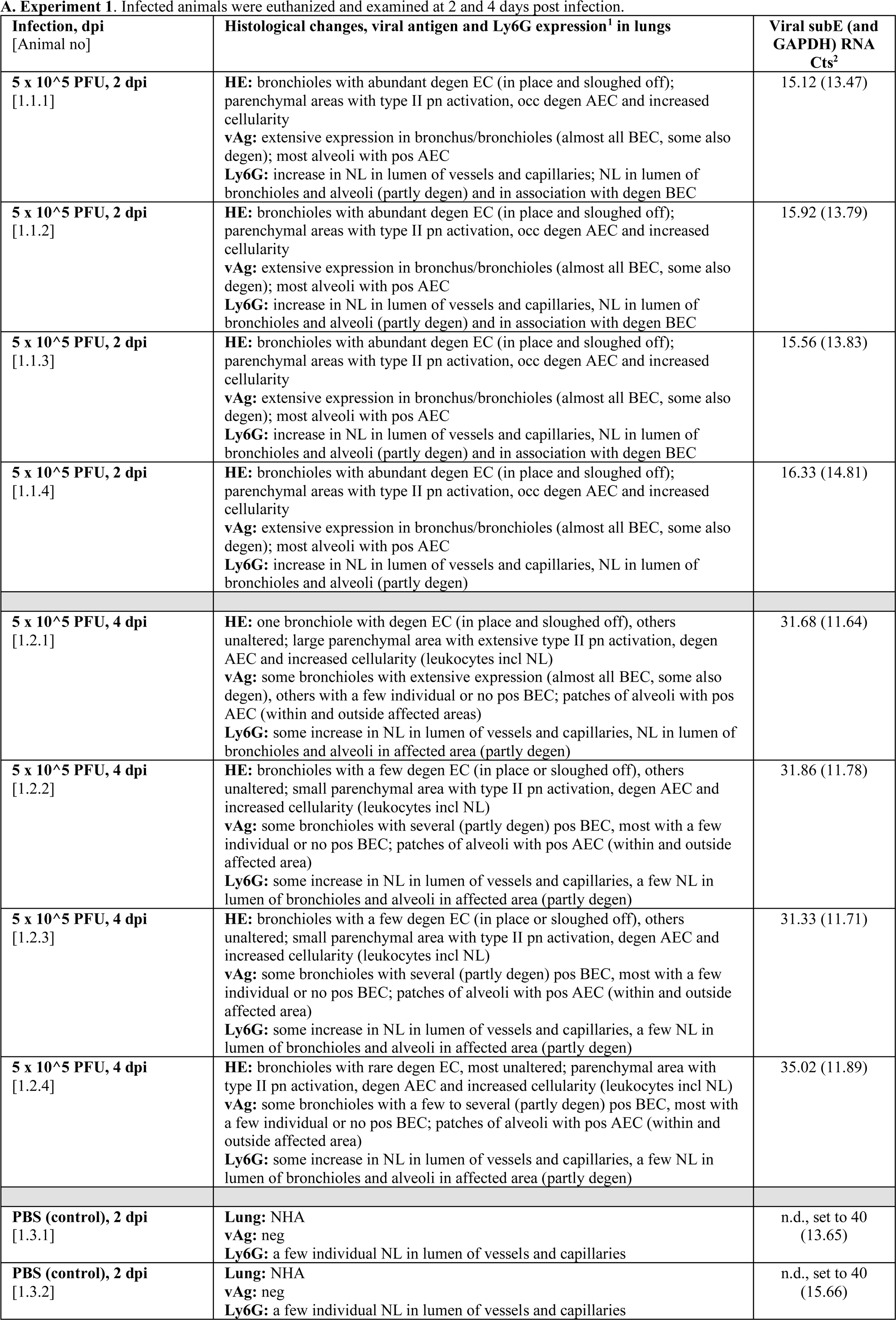
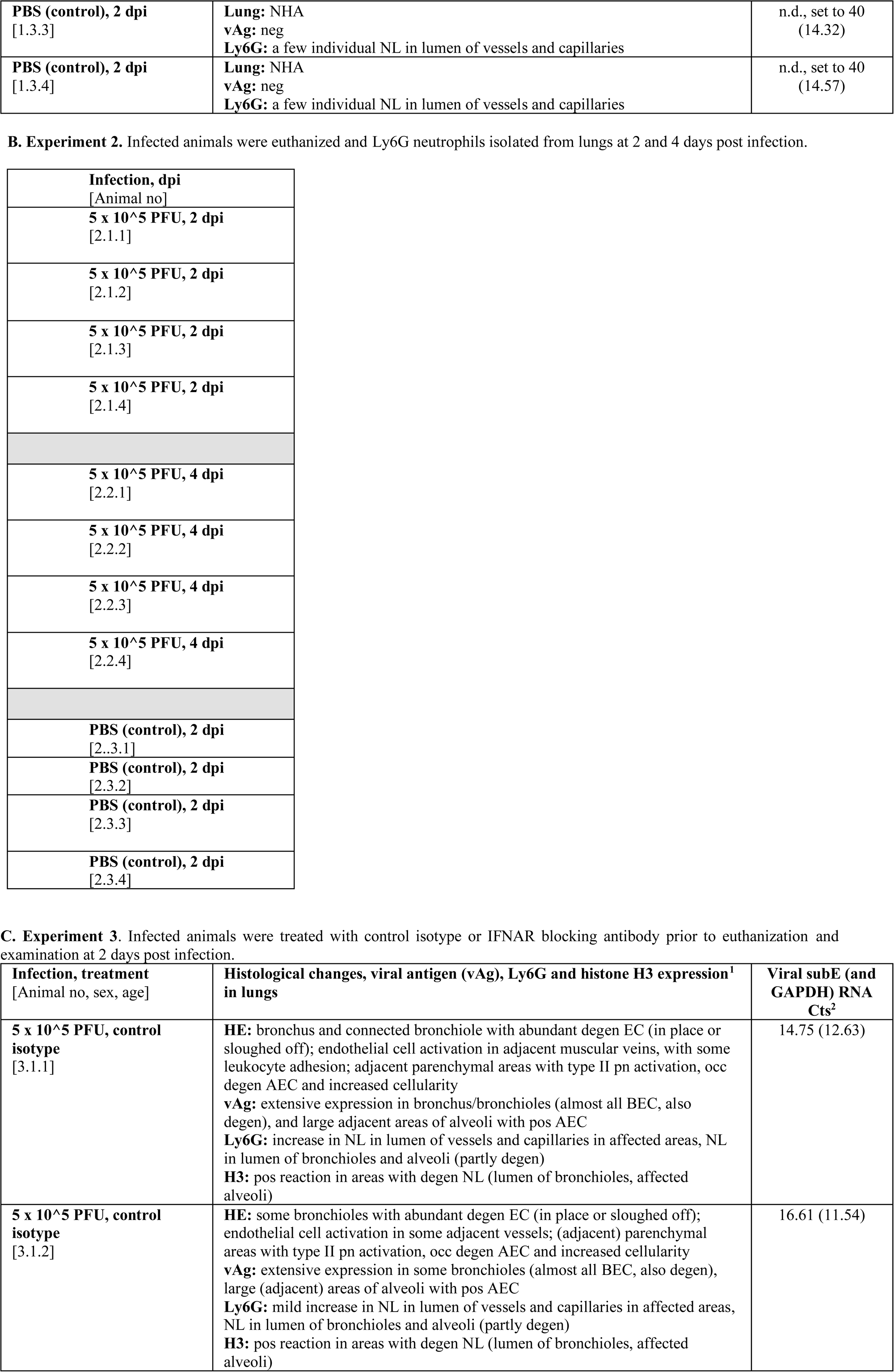
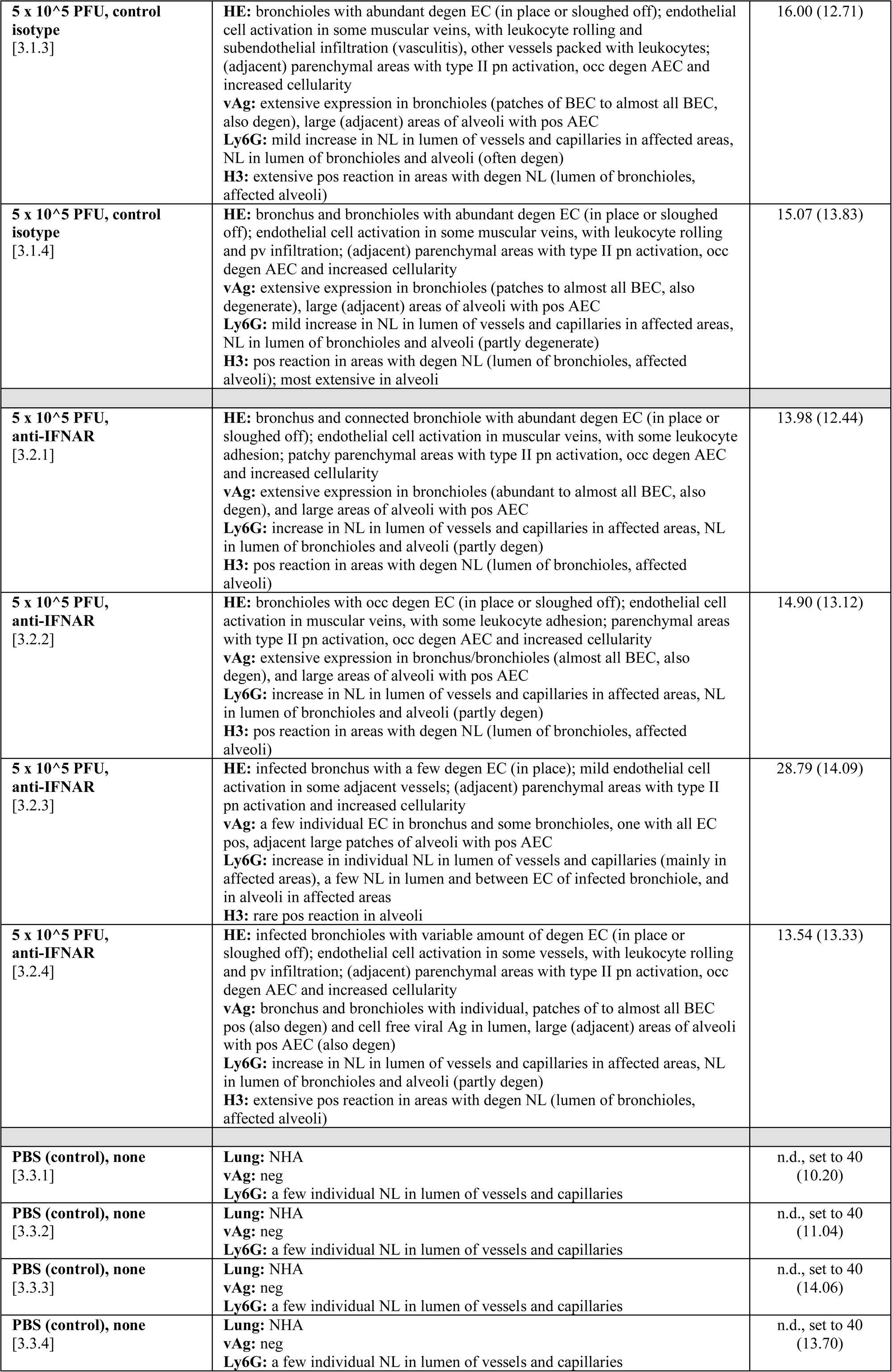
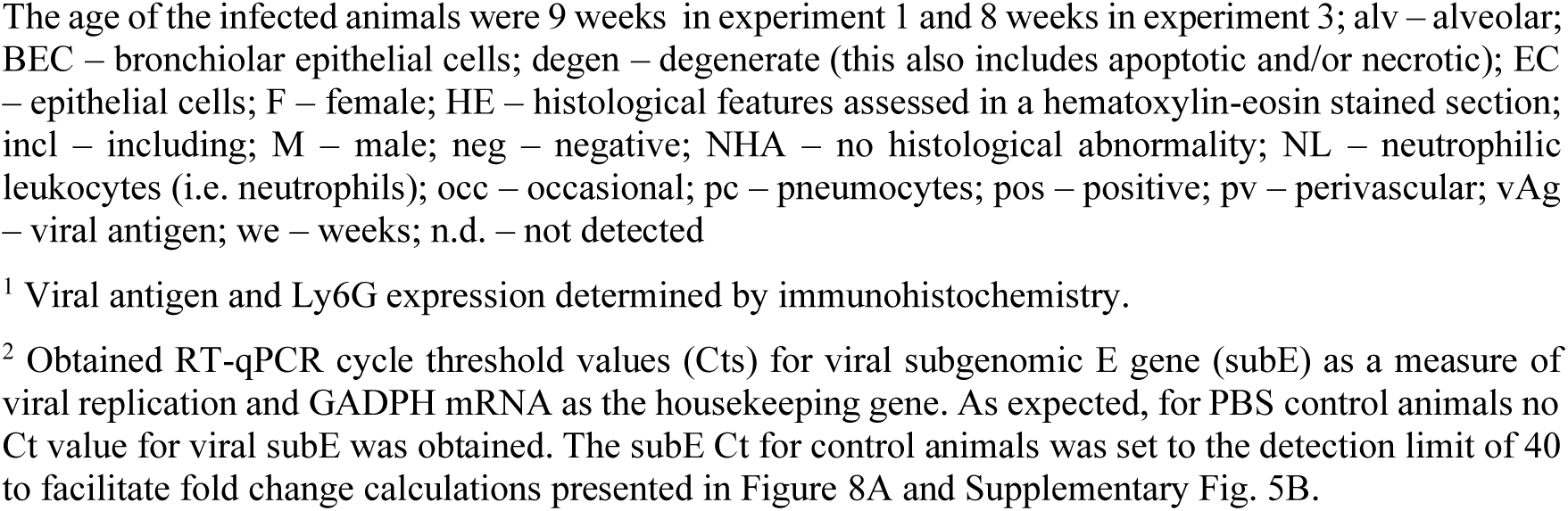
Histological changes as well as SARS-CoV-2 nucleoprotein and RNA expression in female BALB/C mice infected with SARS-CoV-2.

## References

1. Wilk AJ, et al. Multi-omic profiling reveals widespread dysregulation of innate immunity and hematopoiesis in COVID-19. Journal of Experimental Medicine. 2021;218(8). 10.1084/jem.20210582.

2. Blanco-Melo D, et al. Imbalanced Host Response to SARS-CoV-2 Drives Development of COVID-19. Cell. 2020;181(5):1036–1045.e9.

3. Hadjadj J, et al. Impaired type I interferon activity and inflammatory responses in severe COVID-19 patients. Science (1979). 2020;369(6504):718–724.

4. Domizio J Di, et al. The cGAS–STING pathway drives type I IFN immunopathology in COVID-19. Nature. 2022;603(7899):145–151.

5. Barnes BJ, et al. Targeting potential drivers of COVID-19: Neutrophil extracellular traps [preprint]. Journal of Experimental Medicine. 2020;217(6). 10.1084/jem.20200652.

6. Radermecker C, et al. Neutrophil extracellular traps infiltrate the lung airway, interstitial, and vascular compartments in severe COVID-19. Journal of Experimental Medicine. 2020;217(12). 10.1084/jem.20201012.

7. Cabrera LE, et al. Characterization of low-density granulocytes in COVID-19. PLoS Pathog. 2021;17(7). 10.1371/journal.ppat.1009721.

8. LaSalle TJ, et al. Longitudinal characterization of circulating neutrophils uncovers phenotypes associated with severity in hospitalized COVID-19 patients. Cell Rep Med. 2022;3(10). 10.1016/j.xcrm.2022.100779.

9. Schulte-Schrepping J, et al. Severe COVID-19 Is Marked by a Dysregulated Myeloid Cell Compartment. Cell. 2020;182(6):1419–1440.e23.

10. Ueda Y, Kondo M, Kelsoe G. Inflammation and the reciprocal production of granulocytes and lymphocytes in bone marrow. Journal of Experimental Medicine. 2005;201(11):1771–1780.

11. Zuo Y, et al. Neutrophil extracellular traps in COVID-19. JCI Insight. 2020;5(11). 10.1172/jci.insight.138999.

12. Potere N, et al. Interleukin-1 and the NLRP3 inflammasome in COVID-19: Pathogenetic and therapeutic implications. [published online ahead of print: 2022]. 10.1016/j.

13. Lechtenberg BC, Mace PD, Riedl SJ. Structural mechanisms in NLR inflammasome signaling [preprint]. Curr Opin Struct Biol. 2014;29:17–25.

14. Aymonnier K, et al. Inflammasome activation in neutrophils of patients with severe COVID-19. Blood Adv. 2022;6(7):2001–2013.

15. Courjon J, et al. Heterogeneous NLRP3 inflammasome signature in circulating myeloid cells as a biomarker of COVID-19 severity. Blood Adv. 2021;5(5):1523–1534.

16. Leal VNC, et al. Severe COVID-19 patients show a dysregulation of the NLRP3 inflammasome in circulating neutrophils. Scand J Immunol. [published online ahead of print: 2022]. 10.1111/sji.13247.

17. Sefik E, et al. Inflammasome activation in infected macrophages drives COVID-19 pathology. Nature. 2022;606(7914):585–593.

18. Gawish R, et al. ACE2 is the critical in vivo receptor for SARS-CoV-2 in a novel COVID-19 mouse model with TNF-and IFNγ-driven immunopathology. Elife. 2022;11. 10.7554/eLife.74623.

19. Rusanen J, et al. A Generic, Scalable, and Rapid Time-Resolved Förster Resonance Energy Transfer-Based Assay for Antigen Detection-SARS-CoV-2 as a Proof of Concept. [published online ahead of print: 2021]. 10.1128/mBio.

20. Liu P, et al. ExpressAnalyst: A unified platform for RNA-sequencing analysis in non-model species. Nat Commun. 2023;14(1):2995.

21. Emig D, et al. AltAnalyze and DomainGraph: Analyzing and visualizing exon expression data. Nucleic Acids Res. 2010;38(SUPPL. 2). 10.1093/nar/gkq405.

22. Babicki S, et al. Heatmapper: web-enabled heat mapping for all. Nucleic Acids Res. 2016;44(1):W147–W153.

23. Steen CB, et al. Profiling cell type abundance and expression in bulk tissues with CIBERSORTx. Methods in Molecular Biology. Humana Press Inc.; 2020:135–157.

24. Li Y, et al. Alterations of specific chromatin conformation affect ATRA-induced leukemia cell differentiation. Cell Death Dis. 2018;9(2). 10.1038/s41419-017-0173-6.

25. CZ CELLxGENE Discover. Chan Zuckerberg Initiative [Internet]. https://cellxgene.cziscience.com/.

26. Corman VM, et al. Detection of 2019 novel coronavirus (2019-nCoV) by real-time RT-PCR. Eurosurveillance. 2020;25(3). 10.2807/1560-7917.ES.2020.25.3.2000045.

27. Dagotto G, et al. Comparison of Subgenomic and Total RNA in SARS-CoV-2-Challenged Rhesus Macaques. [published online ahead of print: 2021]. 10.1128/JVI.

28. Kant R, et al. Common laboratory mice are susceptible to infection with the SARS-CoV-2 beta variant. Viruses. 2021;13(11). 10.3390/v13112263.

29. Schmid AS, et al. Antibody-based targeted delivery of interleukin-4 synergizes with dexamethasone for the reduction of inflammation in arthritis. Rheumatology (United Kingdom). 2018;57(4):748–755. Doi: 10.1093/rheumatology/kex447.

30. Seehusen F, et al. Neuroinvasion and Neurotropism by SARS-CoV-2 Variants in the K18-hACE2 Mouse. Viruses. 2022;14(5). 10.3390/v14051020.

31. Rosales C. Neutrophil: A cell with many roles in inflammation or several cell types? [preprint]. Front Physiol. 2018;9(FEB). 10.3389/fphys.2018.00113.

32. Walle L Vande, Lamkanfi M. Inflammasomes: Caspase-1-activating platforms with critical roles in host defense. Front Microbiol. 2011;2(JAN). 10.3389/fmicb.2011.00003.

33. Perregaux D, Gabel CA. Interleukin-1β maturation and release in response to ATP and nigericin. Evidence that potassium depletion mediated by these agents is a necessary and common feature of their activity. Journal of Biological Chemistry. 1994;269(21):15195–15203.

34. Gong YN, et al. Chemical probing reveals insights into the signaling mechanism of inflammasome activation. Cell Res. 2010;20(12):1289–1305.

35. Silvin A, et al. Elevated Calprotectin and Abnormal Myeloid Cell Subsets Discriminate Severe from Mild COVID-19. Cell. 2020;182(6):1401–1418.e18.

36. Simard JC, et al. S100A8 and S100A9 induce cytokine expression and regulate the NLRP3 inflammasome via ROS-dependent activation of NF-κB(1.). PLoS One. 2013;8(8). 10.1371/journal.pone.0072138.

37. Jacob C, et al. DMSO-treated HL60 cells: a model of neutrophil-like cells mainly expressing PDE4B subtype. Int Immunopharmacol. 2002;2(12):1647–1656.

38. Tang Y, et al. Excessive IL-10 and IL-18 trigger hemophagocytic lymphohistiocytosis–like hyperinflammation and enhanced myelopoiesis. Journal of Allergy and Clinical Immunology. 2022;150(5):1154–1167.

39. Xue Y, et al. Cardiopulmonary Injury in the Syrian Hamster Model of COVID-19. Viruses. 2022;14(7). 10.3390/v14071403.

40. Vargas F, et al. Intravital imaging of 3 different microvascular beds in SARS-CoV-2-infected mice. Blood Adv. 2023;7(15):4170–4181.

41. Winkler ES, et al. SARS-CoV-2 infection of human ACE2-transgenic mice causes severe lung inflammation and impaired function. Nat Immunol. 2020;21(11):1327–1335.

42. Zhang Y, et al. Neutrophil subsets and their differential roles in viral respiratory diseases [preprint]. J Leukoc Biol. 2022;111(6):1159–1173.

43. Camp J V., Jonsson CB. A role for neutrophils in viral respiratory disease [preprint]. Front Immunol. 2017;8(MAY). 10.3389/fimmu.2017.00550.

44. McKenna E, et al. Neutrophils in COVID-19: Not Innocent Bystanders [preprint]. Front Immunol. 2022;13. 10.3389/fimmu.2022.864387.

45. Lee JS, et al. Immunophenotyping of covid-19 and influenza highlights the role of type i interferons in development of severe covid-19. Sci Immunol. 2020;5(49). 10.1126/sciimmunol.abd1554.

46. Sinha S, et al. Dexamethasone modulates immature neutrophils and interferon programming in severe COVID-19. Nat Med. 2022;28(1):201–211.

47. Takeuchi O, Akira S. Pattern Recognition Receptors and Inflammation [preprint]. Cell. 2010;140(6):805–820.

48. Israelow B, et al. Mouse model of SARS-CoV-2 reveals inflammatory role of type i interferon signaling. Journal of Experimental Medicine. 2020;217(12). 10.1084/JEM.20201241.

49. Broz P, Dixit VM. Inflammasomes: Mechanism of assembly, regulation and signalling [preprint]. Nat Rev Immunol. 2016;16(7):407–420.

50. Hoang TN, et al. Modulation of type I interferon responses potently inhibits SARS-CoV-2 replication and inflammation in rhesus macaques. 10.1101/2022.10.21.512606.

51. Lucas C, et al. Longitudinal analyses reveal immunological misfiring in severe COVID-19. Nature. 2020;584(7821):463–469.

52. Huang C, et al. Clinical features of patients infected with 2019 novel coronavirus in Wuhan, China. The Lancet. 2020;395(10223):497–506.

53. Song X, et al. Little to no expression of angiotensin-converting enzyme-2 on most human peripheral blood immune cells but highly expressed on tissue macrophages. Cytometry Part A. [published online ahead of print: February 1, 2020]. 10.1002/cyto.a.24285.

54. Nikitina E, et al. Monocytes and macrophages as viral targets and reservoirs [preprint]. Int J Mol Sci. 2018;19(9). 10.3390/ijms19092821.

55. Patel AA, Ginhoux F, Yona S. Monocytes, macrophages, dendritic cells and neutrophils: an update on lifespan kinetics in health and disease [preprint]. Immunology. 2021;163(3):250–261.

56. Zhou J, et al. Opsonization of malaria-infected erythrocytes activates the inflammasome and enhances inflammatory cytokine secretion by human macrophages. Malar J. 2012;11. 10.1186/1475-2875-11-343.

57. Labzin LI, Lauterbach MAR, Latz E. Interferons and inflammasomes: Cooperation and counterregulation in disease [preprint]. Journal of Allergy and Clinical Immunology. 2016;138(1):37–46.

58. Pothlichet J, et al. Type I IFN Triggers RIG-I/TLR3/NLRP3-dependent Inflammasome Activation in Influenza A Virus Infected Cells. PLoS Pathog. 2013;9(4). 10.1371/journal.ppat.1003256.

59. Kopitar-Jerala N. The role of interferons in inflammation and inflammasome activation [preprint]. Front Immunol. 2017;8(JUL). 10.3389/fimmu.2017.00873.

60. Rashid F, et al. Roles and functions of SARS-CoV-2 proteins in host immune evasion [preprint]. Front Immunol. 2022;13. 10.3389/fimmu.2022.940756.

61. da Silva RP, et al. Circulating Type I Interferon Levels and COVID-19 Severity: A Systematic Review and Meta-Analysis. Front Immunol. 2021;12. 10.3389/fimmu.2021.657363.

62. Smith N, et al. Defective activation and regulation of type I interferon immunity is associated with increasing COVID-19 severity. Nat Commun. 2022;13(1). 10.1038/s41467-022-34895-1.

63. Robertson SE, et al. Expression and alternative processing of IL-18 in human neutrophils. Eur J Immunol. 2006;36(3):722–731.

64. Mangan MSJ, et al. Targeting the NLRP3 inflammasome in inflammatory diseases [preprint]. Nat Rev Drug Discov. 2018;17(8):588–606.

65. Rubio-Rivas M, et al. WHO Ordinal Scale and Inflammation Risk Categories in COVID-19. Comparative Study of the Severity Scales. J Gen Intern Med. 2022;37(8):1980–1987.

